# Selection of patient-reported outcome measures in pulmonary arterial hypertension clinical trials: a systematic review, meta-analysis and health-related quality of life framework

**DOI:** 10.1101/2024.08.09.24311740

**Authors:** Frances Varian, Rebecca Burney, Charlotte Pearson, Ze Ming Goh, Joseph Newman, Gregg Rawlings, Hamza Zafar, David G Kiely, AA Roger Thompson, Robin Condliffe, Mark Toshner, Ciara McCormack, Iain Armstrong, Tessa Peasgood, Jill Carlton, Alex Rothman

## Abstract

**Introduction:** Health-related quality of life (HRQoL) in pulmonary arterial hypertension (PAH) is valued as an outcome measure by patients, clinicians and regulators. The selection of PROMs for measurement of HRQoL in PAH clinical trials lacks systematic evaluation of their suitability, accuracy and reliability.

**Method:** We report a systematic review (PROSPERO ID: CRD42024484021) following PRISMA guidelines of PROMs selected in PAH clinical trials. PROM measurement properties were then evaluated according to the ten-step COnsensus-based Standards for the selection of health Measurement INstruments (COSMIN) checklist and graded by recommendation for use. Finally, HRQoL was modelled into a conceptual framework using patient interviews and surveys.

**Results:** Screening of 896 records identified 90 RCTs. 43 trials selected PROMs of which 20 were sufficiently validated to detect meaningful change. Of these, 8 trials were adequately powered, using either EQ-5D-5L, SF-36 or the Living with Pulmonary Hypertension Questionnaire (LPHQ). COSMIN evaluation recommended EmPHasis-10 and LPHQ for use (Grade A), however, SF-36 and EQ-5D-5L require further study (Grade B). A conceptual framework of HRQoL was developed from literature comprising 8,045 patients. This framework can be used to visualise the different HRQoL concepts measured by different PROMs.

**Conclusion:** To improve patient-centred research, greater consistency in PROM selection is required. 3 of 90 RCTs have selected COSMIN-recommended PROMs. Whilst the PROMs evaluated require development across the ten areas of psychometric property measurement, EmPHasis-10 and LPHQ can be recommended for use. The ratified conceptual framework can further support PROM selection by identifying the HRQoL concepts they are likely to capture.

**Graphical abstract: selection of patient-reported outcome measures in pulmonary arterial hypertension clinical trials:** COSMIN COnsensus-based standards for the Selection of health-Measurement INstruments, EQ-5D-5L EuroQol-5D-5L; HRQoL health-related quality of life; LPHQ Living with Pulmonary Hypertension Questionnaire, MCID minimal clinically important difference; PAH pulmonary arterial hypertension; PROM patient reported outcome measure, QALY quality-adjusted life years, RCT randomised controlled trial, SF-36 36-item Short Form survey. https://BioRender.com/u80X854

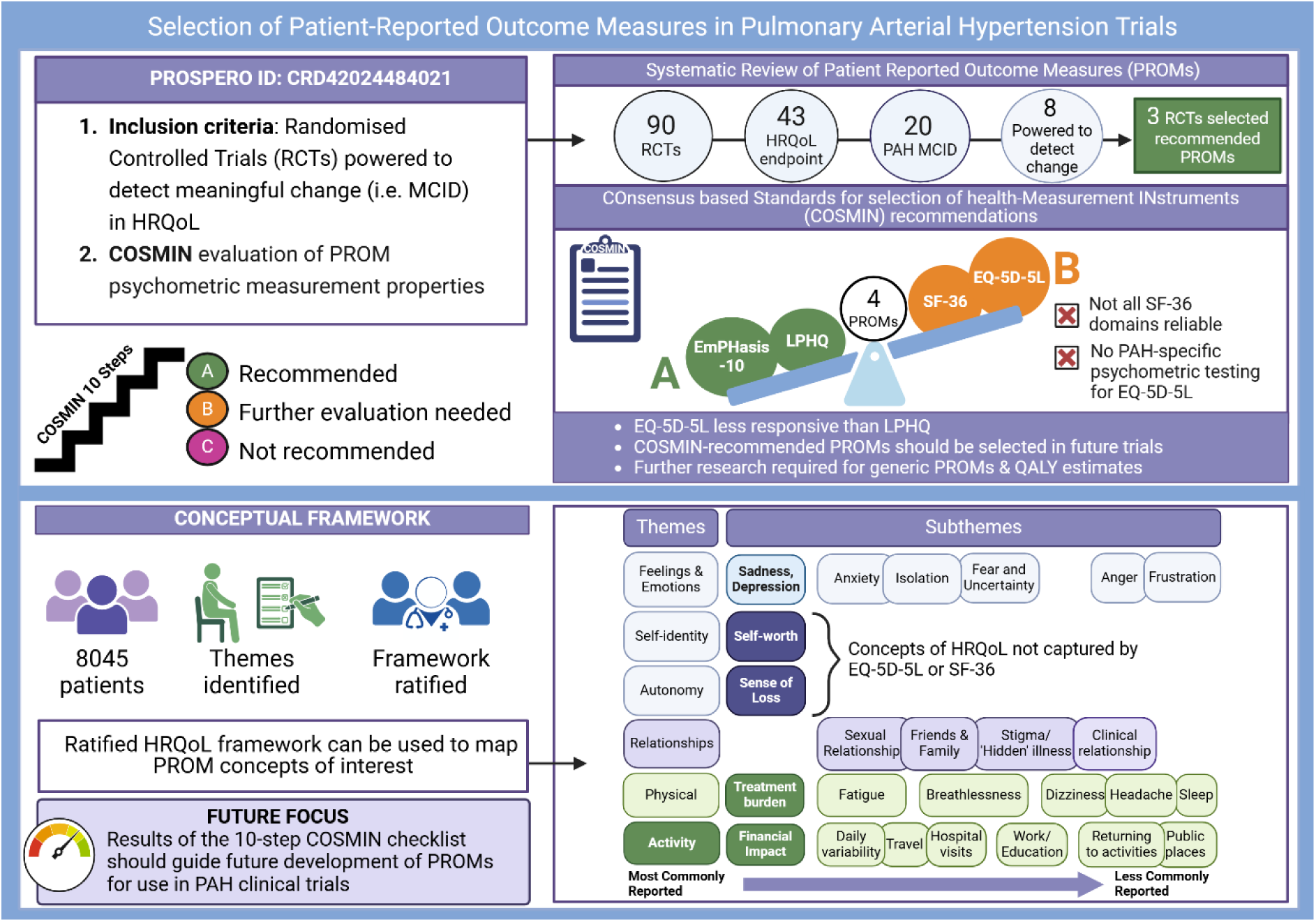

**Plain language summary:** Individuals living with pulmonary hypertension want to know which treatments improve their quality of life related to their health. We use questionnaires to capture the experiences of people living with pulmonary hypertension. Examples of this used in clinical practice are EmPHasis-10. We reviewed all the clinical trials in pulmonary hypertension to see which questionnaires were used to measure health-related quality of life. Some questionnaires may be better at capturing the experience of living with pulmonary hypertension than others. We found 20 clinical trials used a questionnaire that could detect a change in health-related quality of life in pulmonary hypertension. However, only 8 trials were designed to detect a significant treatment impact. We then evaluated these questionnaires against current best practice guidelines to ensure they are fit for purpose. EmPHasis-10 and the Living with Pulmonary Hypertension Questionnaire are preferred from the four evaluated in this study. The final part of this study was to look at what quality of life means for those living with pulmonary hypertension. Data from 8045 patients across the world was used to draft a health-related quality of life framework. We then finalised this design with professionals and patients. This framework can be used in the future to help understand how the well a questionnaire captures things important to those with lived experience of pulmonary hypertension. This will help us to better understand treatments that improve quality of life for people living with pulmonary hypertension.

## Background

Endpoints in randomised controlled trials (RCT) have traditionally focussed on physiological measures including functional markers such as 6-minute walk distance (6MWD).(1,2) However, approaches prioritising clinician-derived endpoints (3–5) can undervalue the patient voice. Patient-reported outcome measures (PROMs) are an instrument developed to capture and quantify the experience of living with a health condition. Improvement in health-related quality of life (HRQoL) is an important treatment goal for clinicians, regulators and patients, yet it is often not examined in clinical trials.(6–11) Furthermore, significant advancements in the diagnosis and treatment of pulmonary arterial hypertension (PAH) mean people are living longer, with a focus not only on length of life, but also quality. There are many challenges in validating PROMs for accurate measurement of HRQoL. A ten-step checklist for PROM risk of bias was developed by the COnsensus-based Standards for the selection of health Measurement INstruments (COSMIN) steering committee.(12) This guidance outlines systematic evaluation of PROMs and facilitates recommendations about PROM suitability.(13,14) This has yet to be undertaken for PROMs in PAH, limiting knowledge of appropriate PROM selection in clinical trials.

PROMs can also be used to evaluate the cost-effectiveness of interventions, typically calculated as Quality Adjusted Life Years (QALYs). To allow such a calculation, PROMs require a value set. Such PROMs are termed a preference-weighted measure (PWM). Value sets are based on the views or preferences of the public and/or patients and vary by country to reflect sociocultural differences.(15,16) A PWM scores each health state described by the PROM as a single value, or ‘utility index’, on a scale, such that 1 represents full health, and zero represents death. A score below zero indicates a health state considered worse than being dead. The index score of a health state can be combined with time spent in that state to estimate QALYs. QALYs are an important outcome for regulatory and clinical decision-making and therefore dependent upon robust PROMs.(17)

PROM selection in clinical trials should follow guidance developed using international Delphi approaches (18) and be evidence-based.(14) Condition-specific PROMs may offer greater sensitivity to changes in HRQoL than generic PROMs, though evidence is limited.(11,19) The condition-specific PROMs for PAH include the Cambridge Pulmonary Hypertension Outcome Review (CAMPHOR), EmPHasis-10, Living with Pulmonary Hypertension Questionnaire (LPHQ) and PAH-SYMptoms and imPACT (PAH-SYMPACT).(20) Guidelines and regulators, such as the US Food and Drug Administration (FDA), recommend that sensitivity to therapeutic change must be interpreted as clinically meaningful (defined as the minimal clinically important difference (MCID)).(11,13,18,21–25) A generic PROM (e.g. SF-36) may be used, providing the instrument has a MCID validated in the population of interest.(18)(14) MCID interpretation comprises one of ten-steps evaluating measurement properties within the COSMIN risk of bias checklist.

We follow the COSMIN systematic review process to facilitate recommendations of which PROMs should be selected in PAH clinical trials (13,14)(13,14). This process can be further enhanced by identifying which HRQoL concepts are captured by the PROM.(26) Developing a conceptual framework aids visualisation of important aspects of HRQoL for people living with pulmonary hypertension.(27,28) The content of PROMs can then be compared to this framework to illustrate which aspects of HRQoL are likely to be measured.

### Aims and Objectives

This is the first systematic review of PROMs selected for use in clinical trials for adults with PAH(20,29–31) to, 1) evaluate appropriate selection with a valid MCID, and 2) compare measurement properties in accordance with the COSMIN checklist, including grading recommendation for use.(12,14,22–24,32). Additionally, to further inform selection of PROMs, we undertake a literature review to develop a conceptual framework which summarises the impact of relevant HRQoL concepts from the perspective of people living with pulmonary hypertension.

## Methods

### Systematic searches

The protocol for the systematic review of PAH RCTs was registered on PROSPERO (CRD42024484021). The COSMIN systematic evaluation is not independently registered. Methodology adhered to the Cochrane Handbook of Systematic Reviews of Interventions and COSMIN guidance.(33) Reporting structure followed the Preferred Reporting Items for Systematic Reviews and Meta-Analyses (PRISMA) statement (see online supplementary figure E1) and PRISMA-COSMIN outcome measurement instruments (online supplementary figure E3 and table E9)(14). MEDLINE (1980 to December 2023) and Cochrane Library (2002 to December 2023) were searched for RCTs evaluating the effectiveness of any clinical intervention in PAH. Inclusion and exclusion criteria are registered on PROSPERO. After removal of duplicates, one author (FV) screened for relevant titles and abstracts before reviewing the full text for eligibility. Where there was uncertainty about the relevance of an article, a second author (JN) reviewed the title and abstract/main text. A third author was available to adjudicate discrepancies. This process was repeated for the PRISMA-COSMIN search strategy and reporting structure (supplementary figure E2 and E3). PRISMA-COSMIN studies included pulmonary hypertension (PH) comprising group 1 and group 4 patients to maximise inclusion of PROM psychometric studies. Forward and backward searches were performed on eligible articles for both searches, and citation searching performed on systematic reviews identified.

### Data Extraction

Five authors (FV, RB, CP, ZMG, JN) extracted information independently from all RCTs using a pre-determined template. This included sample and trial characteristics, primary and secondary outcome measures and results, and details of PROMs used.(34)

### Risk of Bias and Strength of Evidence

Two authors (ZMG, RB) assessed the systematic review risk of bias using the Cochrane RoB2 Toolkit, and strength of evidence according to the Grading of Recommendations Assessment Development Evaluation (GRADE) criteria. The ten-step COSMIN risk of bias checklist was completed by two authors (FV & CP).(32,33) Any disagreements were discussed until consensus was reached. A COSMIN summary of the overall strength of PROM recommendation is made by grading into one of three categories: (A) PROM can be trusted for use with sufficient evidence of psychometric properties; (B) PROM has potential to be recommended for use but insufficient to meet A or C categories; (C) PROMs with high-quality evidence that a measurement property is insufficient and therefore should not be recommended for use.(12,13,35). A description of psychometric terms is available in supplementary table E5.

### Data Analysis

It is recommended that the MCID is considered for sample size calculations for PROM selection.(22–24) To determine whether trials were sufficiently powered for the chosen PROM, the MCID for each instrument was obtained (supplementary table E1). If data were unavailable specifically for PH, an estimated MCID was searched for (1) respiratory conditions and (2) heart failure to maximise PROM inclusion.(29,36–41) GPower v3.1 was used to estimate sample size calculations using the MCID from a two-independent means model for 80% power, 5% significance, one-tailed test. Trials insufficiently powered to detect a meaningful change were excluded.(42,43) Meta-analysis was undertaken per therapy and by PROM in SPSSv28.1.(44)

### Conceptual framework and patient and public involvement and engagement (PPIE)

A scoping literature review was conducted independently by two authors (FV & RB) to identify HRQoL concepts in PH.(24,25,45). Published studies using primary and secondary analytical methods and grey literature, such as surveys asked by Pulmonary Hypertension Associations, were included (supplementary table E6).(6–9,46–54) Seven major themes from an a priori model (27,55) were used to inform the conceptual framework. Subthemes were then extracted and grouped.(27,55) Final subthemes were weighted from most to least commonly reported. Key professional stakeholders from centres in the UK and Ireland then ratified the framework followed by PPIE obtained from representatives from Pulmonary Hypertension Association UK (PHA UK) and patient volunteers registered within Sheffield’s local PPIE PAH network.

Participation was entirely voluntary without reimbursement. The conceptual framework was then used to evaluate the PROMs identified from the COSMIN review in a process called ‘mapping’, allowing visualisation of which HRQoL concepts are captured by which PROM.

## Results

### Systematic review of PROMs selected in PAH clinical trials

The systematic search identified 896 unique records. After screening, 178 remained for full-text review. Overall, 90 RCTs were identified and 43 used PROMs as a secondary endpoint (supplementary table E2). All studies showed some risk of bias (supplementary table E3). The strength of all studies using PROMs was ‘moderate’ (supplementary table E4). There was no reported patient involvement in selection of PROMs in any RCT (supplementary table E2).(18)

MCIDs are available for three of four PH-specific PROMs: EmPHasis-10, LPHQ and CAMPHOR.(29,36,56,57) The MCID for PAH-SYMPACT was excluded as this was in abstract form only with no reported standard deviation.(41,58,59) All available MCID values and methods of derivation are included in supplementary table E1. Figure 1 shows that 20 of the 43 RCTs selected a PROM with an MCID for PAH. Of these, only 8 trials met the full inclusion criteria with adequate power to detect a meaningful change in HRQoL (Table 1).(60–103) PROMs meeting final inclusion (Table 1) were SF-36, EQ-5D-5L, LPHQ and Minnesota Living with Heart Failure-PH (MLWHF-PH). A utility index is available for use of EQ-5D-5L as a PWM for the PAH population(3), but not a specific MCID. The MCID from an interstitial lung disease population was therefore used as a surrogate estimation for sample size to maximise inclusivity.(104)

**Figure 1:**
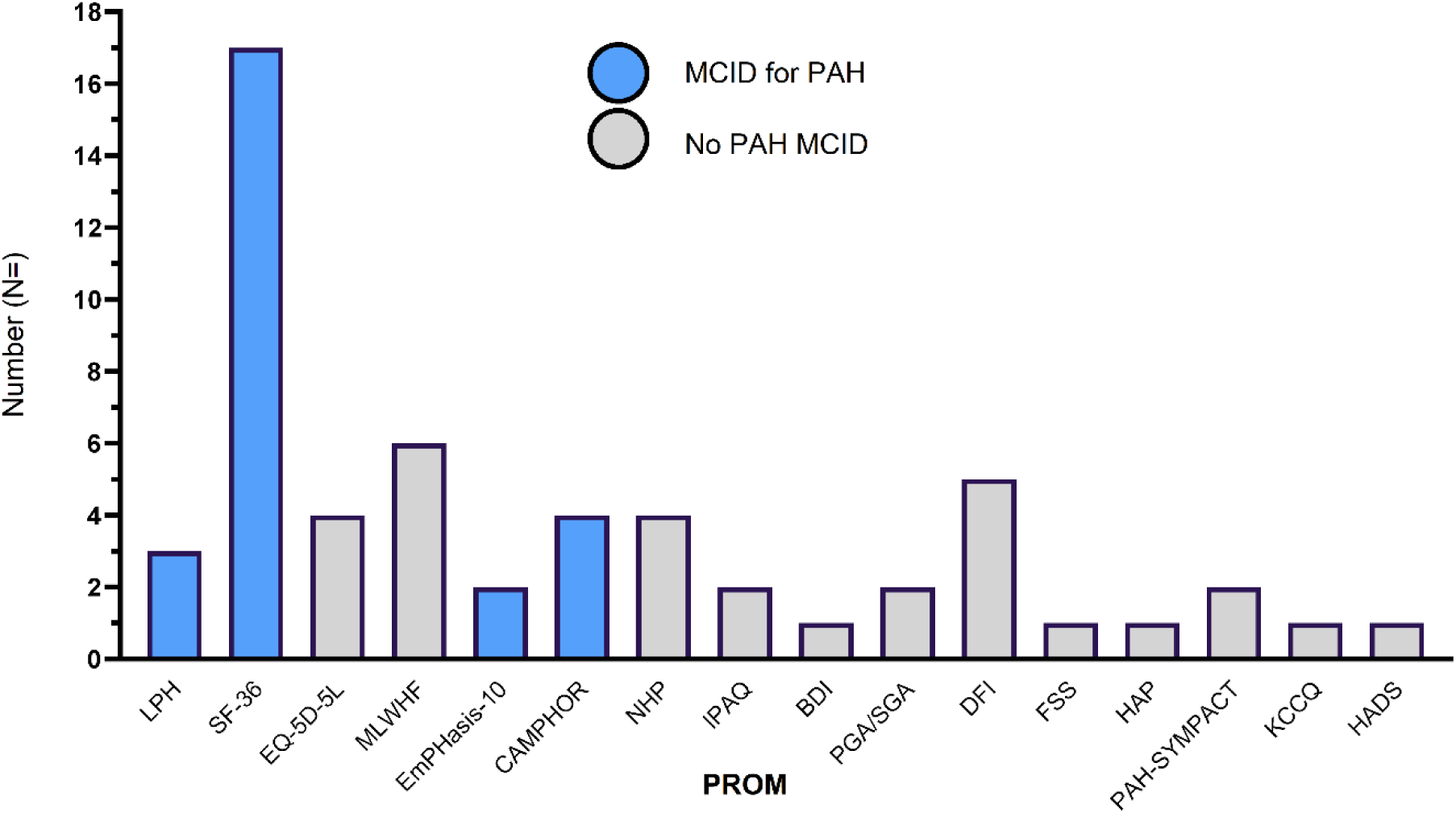
HRQoL Instruments in PAH RCTs from systematic review categorised by ability to distinguish meaningful change in the PAH population. 20 of 43 trials selected an instrument with a MCID for HRQoL. 56 total instruments are included as 13 trials included more than one instrument, see supplementary table E2 for all RCTs with a HRQoL endpoint. No trials reported results in the context of MCID. PAH-SYMPACT MCID evaluation is underway.(59) BDI Beck’s depression inventory, CAMPHOR Cambridge Pulmonary Hypertension Outcome Review, E10 EmPHasis-10, EuroQol(EQ)-5D-5L, DFI dyspnoea fatigue index, FSS fatigue severity score, HADS hospital anxiety and depression questionnaire, HAP human activity profile, IPAQ International Physical Activity Questionnaire, KCCQ Kansas City Cardiomyopathy Questionnaire, LPHQ Living with Pulmonary Hypertension Questionnaire, MCID minimal clinically important difference. MLWHF Minnesota Living with Heart Failure, NHP Nottingham Health Profile, PAH-SYMPACT pulmonary arterial hypertension symptoms and impact questionnaire, PGA patient global assessment, SF-36 Short-Form-36, SGA subject global assessment.

**Table 1.** Characteristics of studies powered for HRQoL 6MWD six-minute walk distance, CPET cardiopulmonary exercise test, CWE clinical worsening events, EQ (EuroQoL)-5D-5L, EQ-VAS EuroQoL visual analogue scale, HRQoL health-related quality of life, NTProBNP N-terminal Pro-Brain Natriuretic Peptide, NYHA New York Heart Failure Association functional score, PROM patient-reported outcome measure, PVR pulmonary vascular resistance, WHO FC World Health Organisation Functional Class ^†^All secondary endpoints.

| Study | N= | Demographics<br>(Age (y), Female (%)) |  | Intervention | PROM<sup>†</sup> | Additional Endpoints | Primary outcome met? |
| --- | --- | --- | --- | --- | --- | --- | --- |
|  |  | Control | Therapy |  |  |  |  |
| EU-TRAIN-01 (2021)(106) | 129 | 55±12.7<br>77.6% | 52.3±12.4<br>69% | Exercise training | SF-36 (all) | Primary 6MWD<br>Secondary WHO FC, CPET | Yes |
| PATENT-2 (2015) (113,159) | 396 | 49±16<br>80% | 49±16<br>80% | Riociguat | LPHQ<br>EQ-5D-5L | Primary 6MWD<br>Secondary CWE, NTProBNP, WHO FC, haemodynamics | Yes |
| PATENT-1 (2013)(160) | 443 | 51±17<br>78% | 51±17<br>79% | Riociguat | LPHQ<br>EQ-5D-5L | Primary 6MWD<br>Secondary CWE, NTProBNP, WHO FC, haemodynamics | Yes |
| TRIUMPH-I (2010)(161) | 235 | 52(18-75)<br>82% | 55(20-75)<br>81% | Inhaled Treprostinil | MLWHF-PH | Primary 6MWD | No |
| EARLY(2008) (162) | 185 | 44±17<br>63% | 45±18<br>76% | Bosentan | SF-36 (all) | Primary 6MWD, PVR<br>Secondary CWE, NTProBNP, haemodynamics | No |
| ARIES2 (2008)(105) | 394 | 51±14<br>68% | 51±15<br>78% | Ambrisentan | SF-36 (physical) | Primary 6MWD<br>Secondary CWE | Yes |
| PACES (2008) (163) | 267 | 48±13<br>77% | 48±13<br>82% | Sildenafil + IV Epoprostenol | SF-36 (all) | Primary 6MWD | No |
| AIR (2002)(164) | 203 | 53±12<br>68% | 51±13<br>68% | Inhaled Iloprost | EQ-5D-5L and EQ-VAS | Primary 6MWD<br>Secondary NYHA, PVR, CWE | Yes |

Bosentan (EARLY)(90)), IV epoprostenol (PACES)(92), and inhaled treprostinil (TRIUMPH-I)(89) did not meet their primary endpoint (6MWD) which was consistent with findings of no improvement in the SF-36 physical functioning domain (Table 1). Significant improvements in 6MWD for ambrisentan (ARIES2)(105) and exercise (EU-TRAIN-01)(106) were reported, however only the MCID was met for the role-physical domain of SF-36 in EU-TRAIN-01(106).

PATENT-1(107) and PATENT-2(108) (riociguat vs placebo) were the only RCTs available for meta-analysis (Figure 2). Two PROMs (EQ-5D-5L and LPHQ) were completed by the same patients. EQ-5D-5L overall appeared less responsive to changes in HRQoL (Cohen’s d ES=0.24, SE=0.08, p<0.001) compared to LPHQ (ES=-0.48, SE=0.11, p<0.001) (Figure 2). Though an exploratory endpoint, it is unclear which country-specific value sets were used for EQ-5D-5L, limiting meaningful interpretation as a PWM.(16,109–112) All trials reported statistical significance (p<0.05) between arms rather than following regulatory recommendations of interpreting results in the context of the MCID. A sustained improvement in HRQoL is shown with all dose-regimes of riociguat compared to placebo, as measured by LPHQ.(108) However EQ-5D-5L was only responsive to change in the 1.5mg subgroup and not the 2.5mg (Figure 2).(113)(108) On further examination, the 1.5mg subgroup had a statistically higher proportion of patients in WHO FC III compared to II (Fisher’s exact p<0.05), supporting the need for evaluation of psychometric property performance. (108)

**Figure 2:**
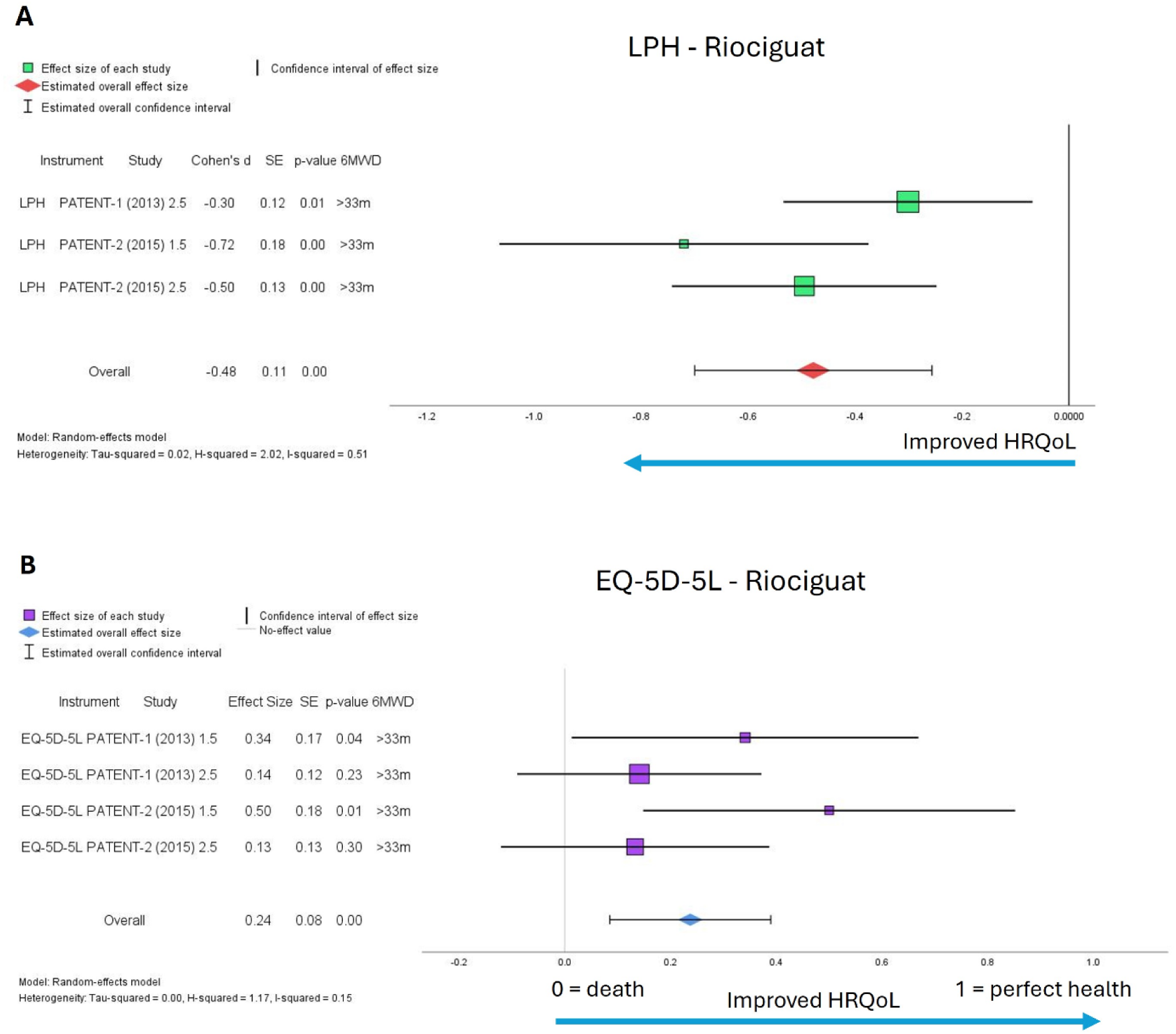
Meta-analysis of HRQoL outcomes for riociguat PROM instruments LPHQ (A) and EuroQoL (EQ)-5D-5L (B). 1.5mg dose in Patent-1 for LPHQ was excluded as subgroup insufficiently powered. HRQoL health-related quality of life, LPHQ Living with Pulmonary Hypertension. Utility index score was not reported with EQ-5D-5L analysis. PROMs delivered at start and week 12 for PATENT-1, and every 2 weeks up to week 8 for PATENT-2 follow-on study. 12-month follow-up data for EQ-5D-5L from PATENT-2 not included. No imputation reported of missing data. *2.5mg riociguat 2013,*(111) *n= 254 (WHO FC III, n = 140 (55%) Vs WHO FC II, n = 108 (43%)** p>0.05; riociguat 1.5mg 2013, n =63 (WHO FC III, n= 39 (62%) Vs WHO FC II, n = 19 (30%)***p<0.0001); 2.5mg riociguat 2015,*(108) *n= 231 (WHO FC III, n= 127 (55%) Vs WHO FC II, n=97 (42%)** p>0.05; 1.5mg riociguat 2015, n = 56 (WHO FC III n= 35(63%) Vs WHO FC II, n =17 (30%)** p<0.005 all Fisher’s exact test*.

For COSMIN evaluation, 369 eligible articles were screened with additional citation searching (n=20) from 3 systematic reviews. EmPHasis-10 was considered relevant for inclusion due to available MCID and selection in two recruiting RCTs with adequate sample size.(114,115) 21 studies demonstrated measurement properties (supplementary figure E3).(20,36,43,56,116–132) MLWHF for Pulmonary Hypertension (MLWHF-PH)(117) was later renamed LPHQ and therefore these instruments are pooled for evaluation.(20,36,116,117) SF-36 is available as both a PWM and PROM. However, the MCID for SF-36 is only available for four of eight domains (physical functioning, role-physical, energy-fatigue and social functioning).(119) EQ-5D-5L is also a PWM with a value-set for PH,(133)(36) though no specific MCID is available and there are few psychometric studies. All PROMs selected from the initial systematic review of clinical trials were included in step ten of the checklist which evaluates PROM responsiveness.(12,13,32,35)

### PROM suitability for the PAH population: reporting the ten-step COSMIN checklist

Steps 1 and 2 of the COSMIN risk of bias assessment involves evaluation of PROM development and consideration of how comprehensively a PROM measures HRQoL in PAH. This is described as content validity. Additional descriptions of COSMIN psychometric terms are in supplementary table E5. COSMIN measurement properties of selected PROMs are outlined in Table 2 with full details in supplementary tables E7 and E8.

**Table 2:**
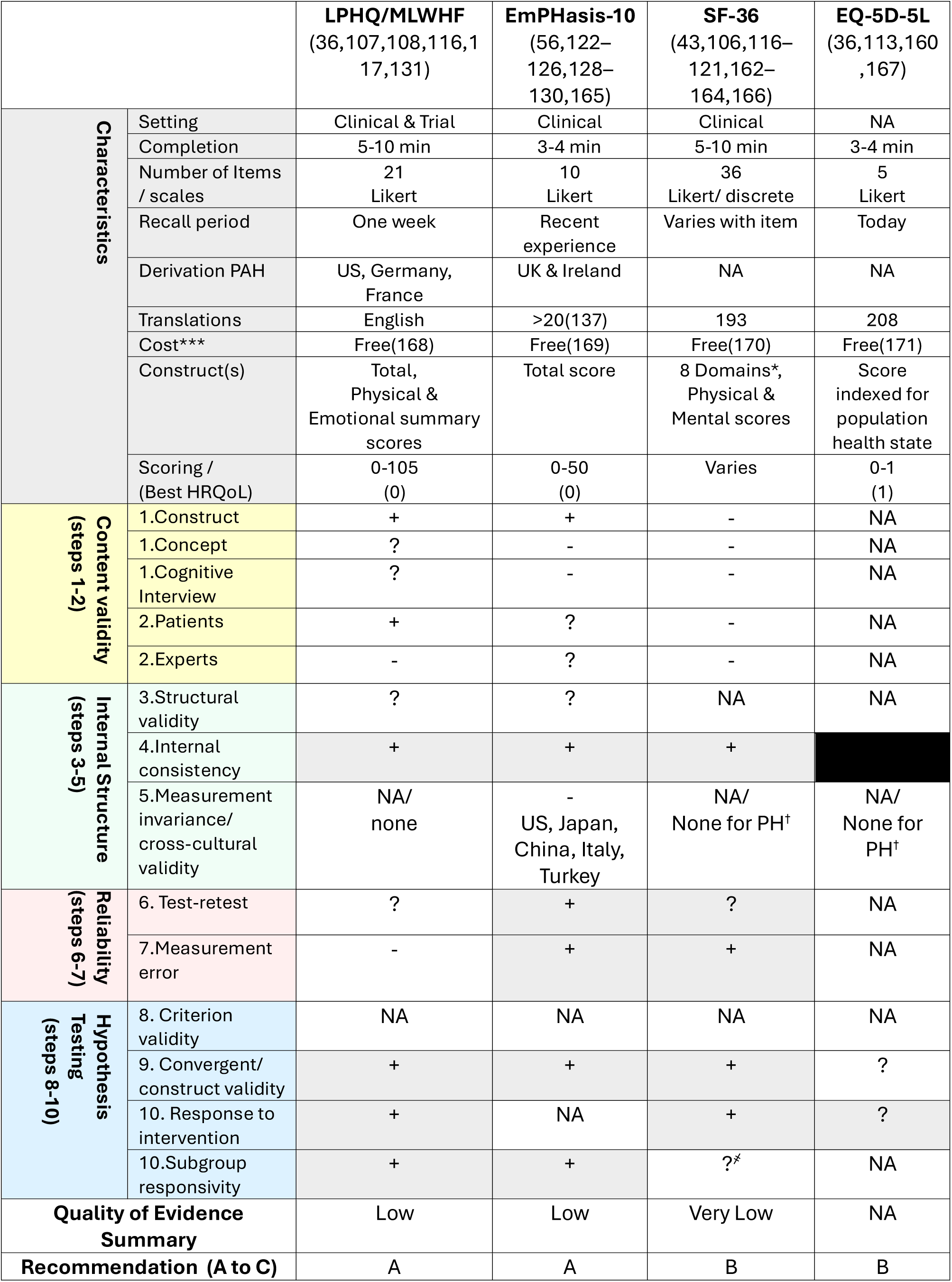
Summary of PROM characteristics, measurement properties and evidence quality sufficient (+), insufficient (–), or indeterminate (?), (NA) not available. Summary of evidence quality based on a modified GRADE approach. Properties with moderate to high evidence quality are shaded grey. Recommendations are made by three categories (A) PROM can be trusted for use with evidence for sufficient content validity and internal consistency, (B) potential to be recommended for use but not categorized as A or C or (C) PROMs with high quality evidence that a measurement property is insufficient and therefore should not be recommended for use. ***non-commercial use, *8 items: P physical, RP role physical, EF energy fatigue, SF social functioning, MH mental health, RE role emotional, GH general health, V vitality. Factor coefficients for mental and physical summary scores are held under copyright, reporting a total overall score is not recommended.(118,172) ^҂^ inconsistencies with item functioning. Full summary of findings available in online supplement tables E7 and E8. ^†^cross-cultural validation may be available for other disease areas/healthy populations however this is outside the scope of this search. If further evaluation is undertaken for measurement invariance in the PAH population, it is recommended to re-evaluate availability cross-culturally. GRADE Grading of Recommendations Assessment Development Evaluation, EuroQol-5D-5L (EQ-5D-5L), LPHQ Living with Pulmonary Hypertension Questionnaire, MLWHF Minnesota Living with Heart Failure, SF Short Form-36

COSMIN recommends that pre- and post-cognitive interviewing with patients and experts must be performed to adequately validate PROM content (steps 1 and 2). LPHQ was the only instrument to satisfy this criteria. No post-hoc cognitive interviewing has been performed in the PAH population for SF-36, EmPHasis-10 or EQ-5D-5L and content validation for these PROMs is therefore insufficient (Table 2).

COSMIN steps 3,4 and 5 outline the appropriate sample sizes and tests in which to evaluate the *internal structure* of the PROM (i.e. how well the questionnaire items perform in measuring HRQoL). The *structural validity (step 3)* should be appropriately analysed and reported as an overall ‘model fit’.(134) The type of model helps determine PROM scoring. LPHQ is a multidimensional model with three scoring methods: physical, emotional and total scores.(36) SF-36 is also multidimensional with eight domains and physical and mental component scores, whereas EmPHasis-10 was derived as a unidimensional structure (i.e. a single, total score).(122) However, a recent model analysis restructured EmPHasis-10 into three scoring components: breathlessness, fatigue and independence.(128) Further clinical evaluation is required. Overall, *structural validity* is not reported for SF-36 or EQ-5D-5L and is inadequate for LPHQ and EmPHasis-10 (Table 2).

Additional checks for internal structure include, step 4 – *internal consistency:* to evaluate whether similar concepts agree, (typically Cronbach’s alpha ≥0.7), and step 5 – *measurement invariance:* stability of PROM responses across different groups (i.e. reducing potential confounders e.g. age, gender). *Internal consistency* is sufficient for LPHQ, EmPHasis-10 and SF-36. This property is not relevant for EQ-5D-5L as items are not inter-related (i.e. they all measure different concepts), with only 1 item per domain (Table 2).(135) No studies adequately considered *measurement invariance* (step 5).(123) Step 5 additionally involves evaluation of *cross-cultural validity*. To use PROMs in multiple different languages and cultures, appropriate statistical testing must be performed.(12,14) While multiple translations are available for EQ-5D-5L and SF-36, there is insufficient validation in the PAH population (Table 2, online supplementary table E8).(136) However, EmPHasis-10 demonstrates strong linguistic testing.(125,126,130,137) Developed in the UK and Ireland, EmPHasis-10 is the only PROM validated in the US, China, Japan, Italy and Turkey.(122,123,125,126,129,130) LPHQ is available in English only, though was developed in the US, France and Germany.

Step 6 evaluates PROM performance under stable conditions, termed *test-retest reliability,* e.g. the intraclass correlation coefficient. This ensures reproducible PROM scores under similar conditions. Reliability should be interpreted within the context of step 7 – *measurement error* e.g. standard error of measurement (SEM). *Measurement error* defines the natural score variation of responders during a period of stability. If the mean variation exceeds the MCID (clinical interpretation of the score), then the PROM becomes invalid, as any detectable change is indistinguishable from normal scoring variation. According to COSMIN, this would raise a major bias concern, rendering the PROM as grade C – not recommended for use.(12,14) Test-retest reliability is sufficient for EmPHasis-10 and indeterminate for LPHQ due to limited evaluation of measurement error (supplementary table E8). Only two SF-36 domains (physical functioning and general health) meet adequate test-retest reliability. Other domains have a high risk of bias due to wide confidence intervals and SEM.(118,119) *Criterion validity* (step 8) can be used to assess sensitivity and specificity of the instrument; however, this requires a ‘gold standard’ measure/PROM and is therefore excluded from this analysis. Step 9 – *Hypothesis testing,* should occur in a stable population and consider 1) how well the PROM correlates with other PROMs developed for PAH (*convergent validity*), 2) how well the PROM discriminates known subgroups e.g. WHO FC (*construct validity*). Correlation with other PAH-specific PROMs is satisfactory for LPHQ, EmPHasis-10 and SF-36, and indeterminate for EQ-5D-5L (supplementary table 8). LPHQ and EmPHasis-10 both show good correlation with WHO FC,(36,116,131) though SF-36 is inadequate.(43,116,119,132) In-hospital invasive haemodynamic assessments have yet to show strong correlation with any PROM.(43,113,114,118–120,124)

Step 10 evaluates responsiveness. Again, described as a *construct approach*, this considers 1) the responsiveness of the PROM with the responsiveness of other PROMs (as exemplified in Figure 2), 2) ability to detect changes in subgroups, and 3) response to an intervention (Table 2, Figure 2). EmPHasis-10 and LPHQ show satisfactory subgroup responsivity (e.g. improvement from WHO FC III to II) (supplementary table E8). EmPHasis-10 is historically absent from RCT interventions, with much-anticipated trials underway.(56,122,123,127).

Summary COSMIN instrument recommendations are grade A for LPHQ and EmPHasis-10 and grade B for SF-36 and EQ-5D-5L. No PROMs received a grade C recommendation. However, the overall quality of evidence for LPHQ and Emphasis-10 is low, and for SF-36 very low (Table 2).

### Developing a HRQoL conceptual framework for PROM content validation

Improving HRQoL matters to people living with pulmonary hypertension. Surveys report this as the most important treatment focus (52-83%) over other outcomes such as life expectancy (33-75%, n=1196, UK, Canada).(10,46) HRQoL concepts of interest may vary between clinical and trial applications,(24,32,45,138) however, recognising their relationship to PROMs is key for appropriate selection. A conceptual framework developed from the Wilson and Cleary – and subsequent – models(27,28,55) was inductively modified to reflect concepts of HRQoL. These subthemes (e.g. ‘stigma’) were identified from combined questionnaires and surveys of 8,045 people living with pulmonary hypertension globally.(19,123,139) Demographics (where available) were reflective of the disease prevalence with a female predominance (79%, n=4,700). Average age of patients was 55 years (range 24-80 years) and 88% self-reported to be Caucasian (supplementary table E6).

Figure 3A summarises the conceptual framework, with six themes and 25 subthemes identified. The framework was ratified by 6 PH consultants, 2 clinical fellows, 1 clinical nurse specialist, 1 physiotherapist, and 1 clinical psychologist, and PPIE obtained from two PHA UK representatives and five patients with relevant demographic representation. No additional themes or subthemes were identified.

**Figure 3:**
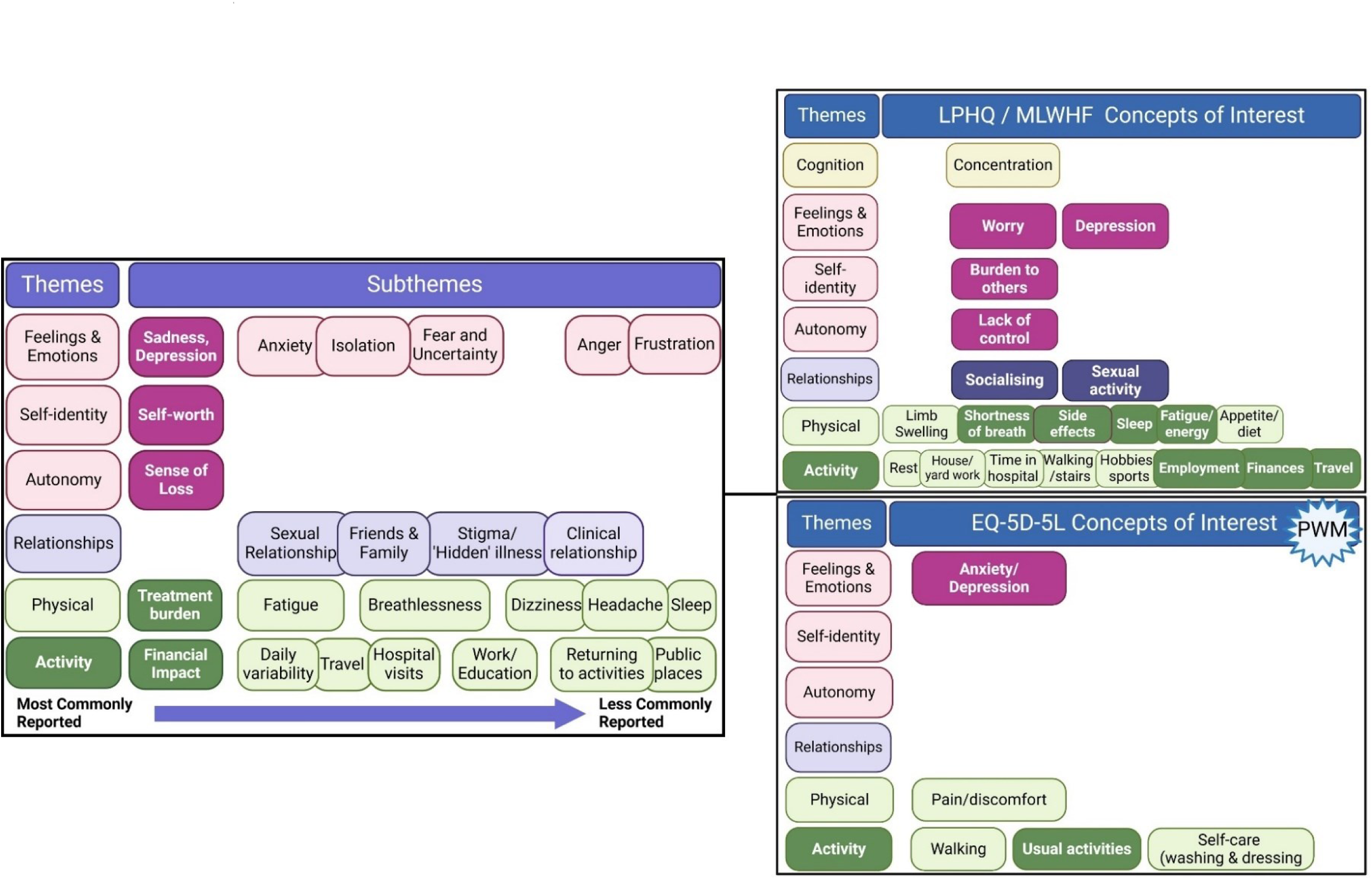
(LEFT) Conceptual framework for PAH HRQoL and (RIGHT) Example of ‘mapping’ of PROMs used in PAH RCTs onto the conceptual framework. [LEFT] Framework of patient-reported themes (n=6) and subthemes (n=25) identified by two independent reviewers on impact of pulmonary hypertension (majority group 1 PAH) on HRQoL. Directly reported concepts are in bold (n= 8045 from supplementary table E6). Concepts may indirectly cross subthemes (cross loading), for example, treatment burden may impact on the EQ-5D-5L item ‘pain/discomfort’ as a reflection of treatment side effects however this is not included within the scope of this analysis. Further PROMs mapped are available in online supplementary figure E4.(114) LPHQ is combined with MLWHF as instruments are identical. [RIGHT] Conceptual mapping of PROMs to HRQoL framework (undertaken by 6 PH consultants, 2 PH clinical fellows, 1 nurse specialist, 1 clinical psychologist, 1 physiotherapist). LPHQ covers all main themes compared to EQ-5D-5L. EQ-5D-5L EuroQol-5D-5L, LPHQ Living with Pulmonary Hypertension Questionnaire, MLWHF Minnesota Living with Heart Failure, PWM preference-weighted measure, SOB shortness of breath, SF-36 Short-Form 36.

The most frequently reported concepts in Figure 3A are presented in bold, with most-to-least common left-to-right and those overlapping having similar weighting. The most reported impacts were activity, sadness/depression, self-worth, sense of loss, treatment- and financial burden. Cultural variation was evident for this latter subtheme and more commonly discussed in surveys and interviews of those living in Canada, USA and China compared the UK and Europe.

PROM content was then ‘mapped’ onto the conceptual framework (Figure 3B) to visualise themes and subthemes likely to be measured. No single PROM covers all subthemes directly. EmPHasis-10 and LPHQ cover all main themes. Two commonly reported themes, self-identity and autonomy, are not specifically captured by EQ-5D-5L or SF-36. In addition, EQ-5D-5L does not capture the impact of PAH on relationships. LPHQ is the only PROM to directly ask about treatment burden by including items on side effects, but may also include items that are less impactful in this patient group (e.g. diet).

## Discussion

This is the first systematic review to evaluate meaningful changes in HRQoL in RCTs in patients with PAH. Based on rigorous methodology using COSMIN guidance, both EmPHasis-10 and LPHQ receive a grade A recommendation for use, whereas SF-36 and EQ-5D-5L receive a grade B recommendation. Of these PROMs EmPHasis-10 provides the broadest scope internationally and is validated on three continents. PROMs meeting COSMIN guidance meet historical FDA recommendations,(11) however this regulatory guidance pre-dates international Delphi consensus.(13,14,18) Whilst SF-36 is the most frequently used PROM in PAH RCTs to date, 6 of 8 SF-36 domains are insufficiently evidenced according to the checklist. No PROMs selected in PAH RCTs to date are adequate for PAH-QALY calculation. All ten areas of the checklist require further study, as the overall quality of evidence summary is low or very low.

To aid future PROM selection and HRQoL evaluation, we developed a conceptual framework which allows visualisation of aspects of HRQoL important to people living with PAH. Six themes and 25 subthemes were identified by researchers and ratified in the conceptual framework.

Whereas both EmPHasis-10 and LPHQ likely capture all major themes, two major themes (self-identity and autonomy) are unlikely to be captured by SF-36 and EQ-5D-5L. Further cognitive interviewing discussing how PROMs relate to this conceptual framework from the patients’ perspective is required.

### Selection of PROMs should be clinically meaningful and patient-centred

PROMs should be resilient to the day-to-day variability in HRQoL. There will be a natural change in score, without a significant change in HRQoL, and this may vary depending on disease severity. Meaningful change in HRQoL, therefore, may not equate to statistical difference.(140,141) It is common practice in PAH to calculate meaningful change thresholds on clinician-derived concepts, or by asking patients to report change using the SF-36 physical functioning domain and compare this to functional changes using 6MWD.(142) Caution is advised when interpreting these MCID values, as they capture a narrow view of HRQoL, as illustrated by multiple concepts of the HRQoL framework.(142,143) PAH-SYMPACT outlined responder thresholds in a recent abstract, calculating the MCID using clinician-derived measures (6MWD, haemodynamics and WHO FC).(59) On review of the COSMIN guidance, this would be considered hypothesis testing/responsiveness (steps 9 and 10), rather than a patient-centred change in HRQoL. CAMPHOR remains the only PROM in PAH to include patient opinion in derivation of a MCID, however the feasibility of using this 65-item questionnaire in clinical trials has potentially limited historical selection.(56,142,144) There is further inaccuracy caused by over-simplifying the MCID at the group-level.(140,145) Multiple MCIDs should ideally be anchored over many individual timepoints to improve sensitivity.(140,142,143) Other factors influencing MCID include direction of change (improvement or deterioration) and individual baseline value, none of which are available for the PAH PROMs evaluated.(145) While highly valuable for trial endpoints, MCIDs should be interpreted with caution, and within the context of measurement error to determine true change over natural responder variability.(21,140,142,145)

### HRQoL is a complex construct to accurately measure

HRQoL is a multifactorial construct with diurnal, daily and lifelong variability. Perception varies across the patient’s lifespan. Changes in values and priorities (*response shift*) depends upon whether patients are ‘pre-diagnosis’, ‘transitioning through diagnosis’ or ‘duration living with PH’. The latter group reportedly face challenges with recognising disease progression and monitoring the condition.(51,146) Registry data shows consistent performance of EmPHasis-10 in patients with recent diagnoses (<6 months) but other time points are lacking, with further research required.(123)

Further complexity is introduced by variations in HRQoL perceptions with age, gender, and disease severity.(147) Age and gender have been shown to influence PROMs.(138,148) These factors require further assessment in the PAH population.(122,139,148,149) Perceptions and response to limitations in activity also vary with individual coping strategies and personality types.(146) Responses may therefore differ depending on the choice of PROM. No PROMs used in PAH trials have specifically addressed variations in activity perceptions in longitudinal subgroup analyses. Consideration of stability and changes across subgroups (steps 9 and 10) is also required (e.g. WHO FC).(140) As shown by the meta-analysis, EQ-5D-5L may be less responsive to changes in WHO FC II compared to WHO FC III, potentially underestimating the HRQoL treatment benefit in this subgroup. Similar responses were shown when comparing EQ-5D-5L with CAMPHOR.(150) Combining PROMs in a trial setting provides useful comparators of responsiveness and is recommended for psychometric evaluation.(12,14)

Development of the conceptual framework helps visualise important HRQoL concepts captured by PROMs. All PROMs capture limitations in activities, however two major themes identified (self-identity and autonomy) are unlikely to be captured by SF-36 and EQ-5D-5L (Figure 3B). While LPHQ has received critique for poor symptom saturation,(29,36) concepts such as ‘time in hospital’ and ‘side effects’ are uniquely captured. Some symptoms may be less relevant. For example, ‘palpitations’, ‘problems with limbs’, and ‘diet’ were not reported as commonly impactful in the HRQoL framework.(6–9,36,46–52) However, LPHQ is also the only instrument to consider financial impact, which may have cultural relevance.(7,9,47,49) It is unclear whether PROMs adequately capture treatment burden (a key subtheme) in PAH, or whether this cross-loads (i.e. is captured elsewhere) with other concepts. Future cognitive interviewing is required to validate all PROMs reviewed. This should consider utilising the conceptual framework to elicit patient interpretation of PROMs, and identifying perceptions of themes and subthemes across the disease course. ‘Mapping’ the content of PROMs to the concepts in this HRQoL framework will also help to solidify PROM content validity.

### Future selection of PROMs in PAH clinical trials

PROMs offer a descriptor for the patient voice, and allow for patient-centred research. This requires appropriate PROM selection with a patient-centred MCID, and prioritisation of PPIE preferences which are reported in line with recommendations.(18,151–156) Greater consistency in PROM selection will improve knowledge of therapeutic outcomes according to lived patient experience. As a minimum, PAH clinical trials should select PROMs with grade A recommendation for use. PROMs may also offer further value in health economic evaluation, though neither generic PWM (EQ-5D-5L and SF-36) can be recommended at this time.(150,157) CAMPHOR(158) is currently the only condition-specific PWM with a value set, however, this is underutilised in RCTs and yet to undergo COSMIN evaluation. Future development of PWMs in PAH should focus on either improving PROMs with a B grade recommendation and/or developing a value set for those with a grade A recommendation. This will support robust evaluation for QALY outcomes.

### Strengths and Limitations

Our systematic review of recent publications was designed with rigour, using multiple reviewers, a minimum of dual coders, and triangulation to enhance quality. Nevertheless, data informing the conceptual framework was not analysed at source and therefore may be subject to bias. However, following UK PPIE opinion, there were no additional concepts added to the framework and, based on the authors experience in international studies in PAH, we consider the framework to be relevant for other countries. As with adaptation of PROMs cross-culturally, future research is required to ensure individual concepts are applicable to the chosen area. This process could offer further understanding of cultural differences in people living with PAH.

Analysis of instrument power was based on MCID; although, as discussed, this may be inadequate, potentially over- or under-estimating the RCTs included.(21) However, this is currently the only available measurement criteria for estimating sufficient responsiveness, and useful for calculating study size.(12,13,35) CAMPHOR has an MCID but did not meet inclusion due to insufficiently powered historical or forthcoming RCTs. As this is currently the only PAH-specific PWM,(150,158) independent COSMIN analysis is warranted. Finally, it is recognised that all PROMs considered in this analysis were developed prior to COSMIN guideline recommendations, and therefore some of the methodological concerns may be overstated due to missing publication details rather than instrument flaws. Despite these challenges and low quality of evidence, two instruments still achieved a grade A recommendation and should be prioritised for selection in future PAH clinical trials.

## Conclusion

Eight PAH clinical trials were adequately powered to detect a meaningful change in HRQoL. Only three of these trials selected PROMs recommended for use. Despite their low grade of evidence, both LPHQ and EmPHasis-10 can be recommended for use in clinical trials. SF-36 and EQ-5D-5L should be used with caution without further examination. Combining these with a grade A recommended PROM may be useful, in addition to offering potential for health economic analyses. The gaps in evidence highlighted using the COSMIN ten-step checklist should be consulted for future psychometric development. These include cognitive interviewing to strengthen content validity, evaluation of natural score variability and further MCID validation, taking into consideration the patient voice, directionality and disease severity.

Selection of PROMs internationally also needs to consider cultural validity, which is not necessarily concordant with language availability. PROM selection can be further supported using the ratified conceptual framework to identify the HRQoL concepts they are likely to capture.

## Contributions

All authors contributed to the development of the manuscript and approved the final version

## Data Availability

All data produced in the present study are available upon reasonable request to the authors

## Acknowledgements

The authors would like to thank the patients from PHA UK and PHA UK representatives who supported the ratification of the conceptual framework. Thank you to Dr Charlie Eliot for ratifying the conceptual framework in addition to the authors.

## Statement of Guarantor

N/A

---

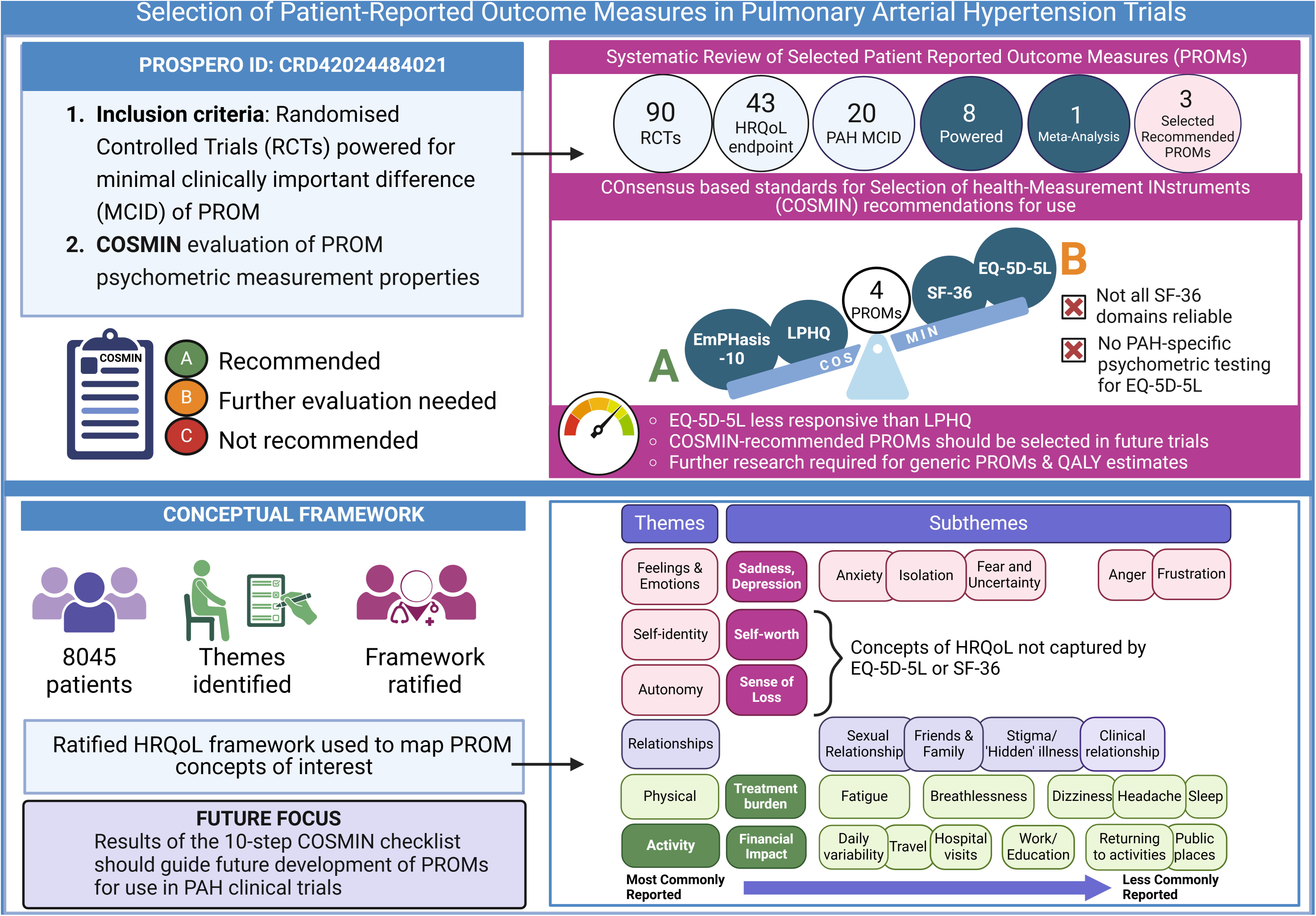

## Notes

### Competing Interest Statement

The authors have declared no competing interest.

### Funding Statement

Wellcome Trust Clinical Research Career Development Fellowship (AMKR: 206632/Z/17/Z), BHF Intermediate Fellowship (AART: FS/18/13/33281), MRC Confidence in Concepts (AMKR), Medtronic External Research Program Award (AMKR), MRC Experimental Medicine grant (AMKR/MT/DGK: MR/W026279/1), BHF Clinical Research Training Fellowship (HZ/AMKR: FS/CRTF/23/24465, MT/JN: FS/CRTF/22/24390). The research was carried out at the National Institute for Health and Care Research (NIHR) Sheffield and Cambridge cardiorespiratory Biomedical Research Centres. AR is grateful to Richard Hughes, whose generous philanthropic support has helped to make this work possible.

### Author Declarations

Systematic review and meta-analysis. Multiple databases were accessed for this review and a secondary literature review of published interviews and surveys. Sources identified are registered on PROSPERO CRD42024484021. All sources are fully references in the manuscript.

### Summary of Updates

Revised title and abstract. Manuscript edited and simplified for the non-PROM audience with emphasis on the 10-step COSMIN checklist.

