## Supplementary tables and figures for "Selection of patient-reported outcome measures in pulmonary arterial hypertension clinical trials: a systematic review, meta-analysis and health-related quality of life framework"

### SUPPLEMENTARY INFORMATION

*Supplementary figure E1: PRISMA flow chart (2020) systematic review*

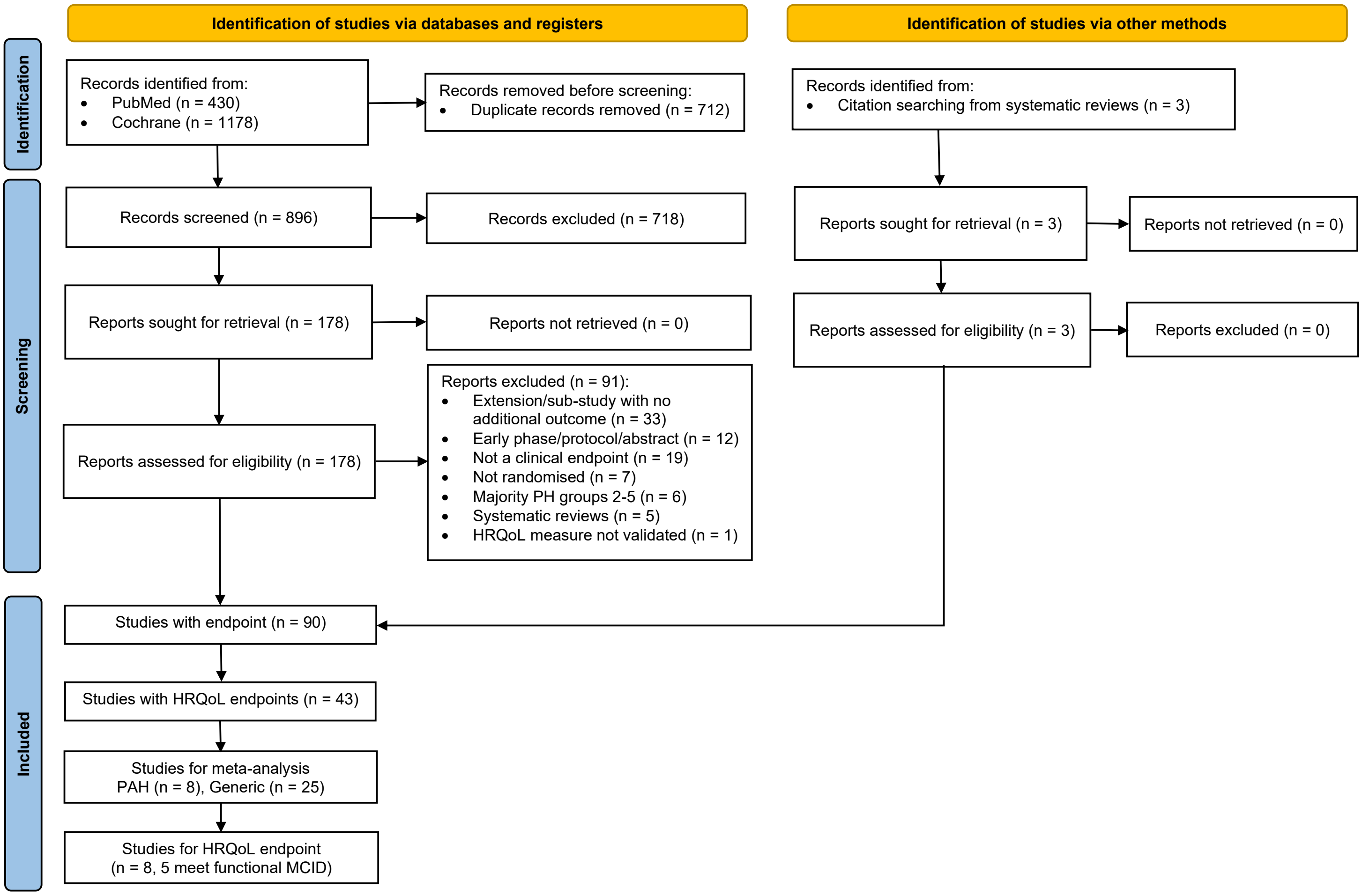

*Supplementary table E1 minimal clinically important differences of HRQoL endpoints.*

Instruments red/grey, no available data for MCID.

| Measure | Derivation | Mean MCID | SD | Distribution method | Disease | Reference |
|---|---|---|---|---|---|---|
| CAMPHOR | Between group | -4 | 6 | Both | PAH | (1) |
| CAMPHOR | Between group | -3 | 6 | Both | PAH | (2) |
| Emphasis-10 | Between group | -7.5 | 11.6 | Both | PAH | (1) |
| EuroQol VAS | Between group | 8 | 11 | Both | COPD | (3) |
| EQ-5D-5L (HSI) | | 0.071 | 0.117 | Both | CAD | (4) |
| EuroQoL (EQ-5D-5L) | | 0.037 | 0.008 | Both | England | (5) |
| EuroQoL (EQ-5D-5L) | | 0.045 | 0.009 | Both | Spain | |
| EQ-5D-5L | | 0.017* | 0.19 | Both | ILD | (6) |
| KCCQ | Individual-responder and between-group | 7 | 18 | Both | HFpEF | (7) |
| LPHQ (total) | Between group | 11 | 22 | Both | PAH | (8) |
| MLWHF | Between group | 8.2 | 26.09 | Both | HF | (9) |
| SF-36 Physical functioning | Between group and individual responder analysis | 13 | | Both p<0.05 for physical functioning only | PAH | (10)¬ |
| SF-36 Role-physical | | 25 | | | PAH | |
| SF-36 Social functioning | | 21 | | | PAH | |
| SF-36 vitality | | 15 | | | PAH | |
| PAH-SYMPACT | Between group | -0.1 to -0.3 | NA | Anchor | PAH | Abstract only (11–13) |
| FSS | | | | | | |
| HAP | | | | | | |
| IPAQ | | | | | | |
| SGA | | | | | | |
| DFI | | | | | | |
| NHP | | | | | | |

Mean MCID and standard deviation reported as between-group for purposes of power calculations.

BDI Beck's depression inventory, CAMPHOR Cambridge Pulmonary Hypertension Outcome Review, E10 EmPHasis-10, EuroQol(EQ)-5D-5L, DFI dyspnoea fatigue index, FSS fatigue severity score, HADS hospital anxiety and depression questionnaire, HAP human activity profile , ILD interstitial lung disease, IPAQ International Physical Activity Questionnaire, IPF idiopathic pulmonary fibrosis, KCCQ Kansas City Cardiomyopathy Questionnaire, LPHQ Living with Pulmonary Hypertension Questionnaire, MCID minimal clinically important difference. MLWHF Minnesota Living with Heart Failure, NHP Nottingham Health Profile, PAH-SYMPACT Pulmonary Arterial Hypertension SYmptoms and iMPACT questionnaire, PGA patient global assessment, SF-36 Short-Form-36, SGA subject global assessment, VAS Visual Analogue Scale

*Range reported in literature depending on which anchor used. Calculation for 6MWD MCID at 35.4m is 0.017 reported anchor method. ¬standard deviation for 6MWD from MCID anchor where available. Where MCID was unavailable, a second author (JN) confirmed absence of MCID using search criteria in PubMed and the PROMs Data Archive: [instrument_name] AND [MCID OR MID OR Minimal].

*Supplementary table E2: HRQoL Endpoints in PAH-RCTs*

| Study (n = 43) | N= | HRQoL Instrument | HRQoL outcome powered for MCID? | Primary Endpoint | Summary of all Endpoints | | | | | | | PPIE? Y/N |
|---|---|---|---|---|---|---|---|---|---|---|---|---|
| | | | | | 6MWD | M&M | WHO FC | NTProBNP | Borg | Haemodynamics | Other(s) | |
| STELLAR 2023(14) | 323 | PAH-SYMPACT | N/A | 6MWD | 0 | 1 | 1 | 1 | 0 | 1 | CPET, Risk | N |
| TRACE 2023(15) | 108 | PAH-SYMPACT | N/A | Actigraphy-assessed DLPA (recorded by using an accelerometer) and total daily physical activity | 0 | Surv 0 | 1 | 0 | 1 | 1 | CPET, CWE | N |
| Kahraman et al (2023)(16) | 31 | IPAQ-SF, NHP | Unlikely | Quadriceps muscle strength | 1 | Surv 0 CWE 0 | 0 | 0 | 0 | 0 | CPET, PRO: Muscle strength | N |
| ARROW(2021)(17) | 151 | SF-36, EmPHasis-10 | N | PVR | 1 | CWE 1 Surv 0 | 1 | 1 | 1 | 1 | | N |
| EU-TRAIN-01(2021)(18) | 129 | SF36 | Y | 6MWD | 1 | 0 | 1 | 0 | 1 | 0 | CPET, PRO | N |
| Hemnes et al (2021)(19) | 42 | SF36, EmPHasis10 | N | Step counts | 1 | CWE 1 Surv 0 | 1 | 0 | 0 | 1 | CTPET | N |
| Howard et al (2021)(20) | 56 | CAMPHOR, PGA | N | CPET | 1 | 0 | 1 | 1 | 1 | 0 | 0 | N |
| PULSAR (2021)(21) | 106 | N | N/A | PVR | 1 | 0 | 0 | 0 | 0 | 1 (PVR) | | N |
| REPLACE (2021)(22) | 226 | N | N/A | Combined (absence CWE, improvement in 2/3 of 6MWD, WHOFC and NTPRoBNP) | 1 | CWE 1 Surv 0 | 0 | 1 | 0 | 0 | | N |
| TRITON (2021)(23) | 247 | N | N/A | PVR | 1 | CWE 1 Surv 0 | 1 | 1 | 0 | 1 | Risk Score | N |
| Aslan et al (2020)(24) | 30 | IPAQ, MLHF | N | MIP/MEP | 1 | 0 | 0 | 0 | 0 | 1 | CTPET: spirometry | N |
| Kahraman et al (2020)(25) | 31 | IPAQ-SF, NHP | Unlikely | Quadriceps muscle strength | 1 | 0 | 0 | 0 | 0 | 0 | PRO: muscle strength | N |
| Yilmaz et al (2020)(26) | 11 | SF36 | N | 6MWD and CPET | 1 | 0 | 1 | 0 | 1 | 0 | CTPET | N |

| | | | | | | | | | | | | |
|---|---|---|---|---|---|---|---|---|---|---|---|---|
| AMBITION (2019)(27) | 500 | N | N/A | Hospitalisation | 0 | 0 | 0 | 0 | 0 | 0 | | N |
| Babu et al (2019)(28) | 84 | SF36 | N | 6MWD | 1 | 0 | 0 | 0 | 0 | 1 | Signs/Sx | N |
| GALILEO-PAH (2017)(29) | 22 | SF36 & LPHQ | N | 6MWD | 1 | 0 | 0 | 0 | 0 | 0 | Signs/Sx | N |
| Han et al (2017)(30) | 27 | MLWH | N | 6MWD | 1 | 0 | 1 | 1 | 0 | 1 | | N |
| FREEDOM-C2 (2013)(31) | 132 | DFI | N/A | 6MWD | 1 | CWE 1 Surv 0 | 1 | 1 | 0 | 0 | Signs/Sx | N |
| Van Campen et al (2016)(32) | 18 | MLHFQ | N | RVEF (CMR) | 1 | 0 | 1 | 1 | 0 | 0 | TTE | N |
| GRIPHON (2015)(33) | 397 | N | N/A | Morbidity and Mortality | 1 | 1 | 1 | 1 (exploratory) | 0 | 0 | | N |
| PATENT-1 (2013)(34) | 443 | LPHQ EQ-5D-5L | Y – 2.5mg dose Y – all doses | 6MWD | 1 | CWE 1 Surv 0 | 1 | 1 | 1 | 1 | | N |
| PATENT-2 (2015)(35,36) | 396 | LPHQ EQ-5D-5L | Y – all doses | 6MWD | 1 | CWE 1 Surv 1 | 1 | 1 | 1 | 0 | | N |
| Saglam et al (2015)(37) | 29 | NHP | | Inspiratory muscle training on functional capacity | 1 | 0 | 0 | 0 | 0 | 0 | Spirometry, fatigue severity scale, mMRC dyspnoea score | N |
| Ulrich et al (2015)(38) | 23 | SF-36, MLHF | N | 6MWD, SF-36 | 1 | 0 | 1 | 1 | 0 | 0 | TTE, sleep, CPET | N |
| Chan et al (2013)(39) | 23 | SF-36, CAMPHOR | | 6MWD | 1 | 0 | 0 | 0 | 0 | 0 | bioimpedence cardiography, CPET | N |
| IMPRES (2013)(40) | 202 | N | N/A | 6MWD | 1 | CWE 1 Surv 0 | 1 | 1 | 0 | 1 | | N |
| Weinstein et al (2013)(41) | 24 | Fatigue severity scale, human activity profile | Unlikely | FSS and HAP | 0 | CWE 1 Sur 0 | 0 | 0 | 0 | 0 | | N |
| FREEDOM-C (2012)(42) | 350 | DFI | N/A | 6MWD | 1 | CWE 1 Surv 0 | 1 | 0 | 1 | 0 | | N |
| ASA-STAT (2011)(43) | 92 | N | N/A | 6MWD | 1 | CWE 1 Surv 0 | 1 | 1 | 0 | 1 | | N |

| | | | | | | | | | | | | |
|---|---|---|---|---|---|---|---|---|---|---|---|---|
| FREEDOM-M (2011)(44) | 349 | DFI | N/A | 6MWD | 1 | 0 | 1 | 0 | 1 | 0 | Signs & Sx | N |
| Ghofrani et al (2010)(45) | 59 | N | N/A | 6MWD | 1 | 0 | 1 | 0 | 0 | 1 | | N |
| TRIUMPH-I (2010)(46) | 235 | MLWHF | Y | 6MWD | 1 | CWE 1 Surv 0 | 1 (NYHA) | 1 | 1 | 0 | | N |
| EARLY(2008)(47) | 185 | SF36 | Y (not V or SF) | PVR & 6MWD | 1 | CWE 1 Surv 0 | 1 | 1 | 1 | 1 | | N |
| ARIES1/2 (2008)(48) | 394 | SF36 | Y | 6MWD | 1 | CWE 1 Surv 0 | 0 | 1 | 1 | 0 | | N |
| PACES (2008)(49) | 267 | SF36 | Y | 6MWD | 1 | CWE 1 Surv 0 | 0 | 0 | 1 | 0 | | N |
| COMBI (2006)(50) | 40 | EQ-5D-5L | Y | 6MWD | 1 | CWE 1 Surv 0 | 1 (NYHA) | 0 | 0 | 0 | CPET | N |
| Mereles et al (2006)(51) | 30 | SF36 | N | 6MWT & SF-36 | 1 | 0 | 1 | 0 | 1 | 1 | TTE | N |
| Galie et al (2005)(52) | 64 | Subject Global Assessment | ?N – not on MCID | 6MWD | 1 | 0 | 1 | 0 | 1 | 1 | | N |
| SERAPH (2005)(53) | 26 | KCCQ | N | CMR | 1 | 0 | 1 | 1 | 1 | 0 | TTE | N |
| BREATHE-2 (2004)(54) | 33 | DFI | N/A | Haemodynamics (TPR) | 1 | 0 | 1 (NYHA) | 0 | 0 | 1 | | N |
| Barst et al (2003)(55) | 116 | MLWHF | N/A | Disease progression | 1 | 0 | 0 | 0 | 1 | 1 | CPET, Signs/Sx | N |
| AIR (2002)(56) | 203 | EQ-5D-5L | Y | NYHA & 6MWD 10% | 1 | 0 | 1 (NYHA) | 0 | 1 (Mahler) | 1 | | N |
| Barst et al (1996)(57) | 20 | Chronic HF, NHP, DFI | N/A | 6MWD | 1 | 0 | 0 | 0 | 0 | 1 | | N |

6MWD six-minute walk distance, CPET cardiopulmonary exercise test, HRQoL health related quality of life, LPHQ living with pulmonary hypertension questionnaire, M&M morbidity and mortality (including clinical worsening events), PVR pulmonary vascular resistance, Surv= survival, WHO FC World Health Organisation Functional Class

*Supplementary table E3: Risk of Bias of Studies with HRQoL PROM*

***Shaded rows (grey) are powered for HRQoL***

| Study | Random Sequence Generation | Allocation Concealment | Blinding | Incomplete Outcome Data | Selective Reporting | Other Bias | Risk of Bias |
|---|---|---|---|---|---|---|---|
| BREATHE-2 2004 | Randomised but not described how | No information provided | Double-blind | Description of withdrawals & method of imputation for missing data | All expected outcomes are reported | Yes<br><br>(2:1 randomisation without justification + Small sample size) | Some concerns |
| SERAPH 2005 | Randomised but not described how | No information provided | Double blind | Description of withdrawals & method of imputation for missing data | All expected outcomes are reported | Small sample size | Some concerns |
| Galie et al<br><br>2005 | Randomised but not described how | No information provided | Double-blind<br><br>(Open-label after 12 weeks – unlikely to affect result) | Description of withdrawals & method of imputation for missing data | All expected outcomes are reported | No | Some concerns |
| ARIES1/2 2008 | Patient randomized stratified by baseline therapy | No information provided | Blinding of participants, key personnel and outcome ensured, unlikely that blinding could have been broken | Description of withdrawals and method of imputation for missing data | All expected outcomes are reported as pre-specified | No | Low |
| EARLY<br><br>2008 | Patients randomly allocated using a computer random number generator | Central allocation via integrated voice recognition system | Blinding of participants, key personnel and outcome ensured, unlikely that blinding could have been broken | Description of withdrawals and method of imputation for missing data | All outcomes are reported | No | Low |

| | | | | | | | |
|---|---|---|---|---|---|---|---|
| PATENT-1 2013 | Patients randomly allocated using a computer random number generator | Central allocation via Interactive Voice Response System | Blinding of participants, key personnel and outcome ensured, unlikely that blinding could have been broken | Description of withdrawals and method of imputation for missing data. Discontinuation balanced between groups | All outcomes are reported. Study protocol and supplementary appendix is available | No | Low |
| PATENT-2 2015 | Randomisation in PATENT-1 | Allocation in PATENT-1 | Double blind | Description of withdrawals and method of imputation for missing data. Discontinuation | All outcomes are reported. | Yes (Open-label, patient withdrawal) | Low |
| van Campen et al 2016 | Randomised but not described how | No information provided | Double-blind | Description of withdrawals and method of imputation for missing data. | All expected outcomes are reported | Yes (Target sample size for power not achieved, small sample size, open-label) | Some concern |
| ARROW 2021 | Randomised with stratification | Central allocation through interactive voice-response or web-response system | Double-blind | Description of withdrawals and method of imputation for missing data | All expected outcomes are reported | Yes (High number of patients on combined therapy, relatively low-risk status) | Low |
| TRACE 2023 | Randomisation through computer generated digits | Allocation via Interactive Response Technology | Double-blind | Description of withdrawals & method of imputation for missing data | All expected outcomes are reported | Yes (Involvement of sponsor) | Low |
| Barst et al 1996 | Computer-generated, adaptive randomization with stratification according to functional class, study center, and baseline vasodilator use | No information provided | No blinding | No description of withdrawals | All expected outcomes are reported | Yes (Patients receiving conventional therapy had better baseline exercise capacity) | Some concern |

| | | | | | | | |
|---|---|---|---|---|---|---|---|
| AIR<br>2002 | Randomised based on after stratification according to NYHA and type of PH | No information provided | No information provided | Withdrawals not described | All expected outcomes are reported | Yes<br>(uncontrolled trial) | Some concern |
| Barst et al 2003 | Patients randomized, not further specified | No information provided | Blinding of participants, key personnel and outcome ensured, unlikely that blinding could have been broken | Description of withdrawals and method of imputation for missing data | All expected outcomes are reported | No | Low |
| COMBI<br>2006 | Patients randomized using pre-prepared, sealed envelopes | Central allocation using pre-prepared sealed envelopes | No blinding but outcome not likely to be influenced by lack of blinding | Description of withdrawals & method of imputation for missing data | All outcomes are reported | Yes<br>(small sample size + Open-label design) | High |
| PACES<br>2008 | Patients randomly allocated using a computer random number generator | Central allocation | Blinding of participants, key personnel and outcome ensured, unlikely that blinding could have been broken | Description of withdrawals & method of imputation for missing data. More lost to follow up in placebo group | All expected outcomes are reported | No | Low |
| TRIUMPH-I 2010 | Patients randomized, not further specified | No information provided | Blinding of participants, key personnel and outcome ensured, unlikely that blinding could have been broken | Description of withdrawals and method of imputation for missing data | All outcomes are reported | No | Low |
| FREEDOM-C 2012 | Patient randomized stratified & blocked by background therapy & baseline 6MWD | No information provided | Blinding of participants, key personnel and outcome ensured, unlikely that blinding | Description of withdrawals and on the method of imputation for missing data, but more | All outcomes are reported | No | Low |

| | | | | | | | |
|---|---|---|---|---|---|---|---|
| | | | could have been broken | discontinuations in treatment group | | | |
| Ulrich et al 2015 | Randomised through computer generated list with balanced block design | No information provided | Double-blind | Description of withdrawals and method of imputation for missing data | All expected outcomes are reported | Yes<br><br>(Small sample size in each group) | Some concern |
| Han et al<br><br>2017 | Randomised but not described how | No information provided | No blinding | Description of withdrawals & method of imputation for missing data | All expected outcomes are reported | Small sample size +<br><br>Open-label | High |
| STELLAR 2023 | Randomised but not described how | No information provided | Double-blind | No description of withdrawals | All expected outcomes are reported however 48% drop-out of PRO data which was then imputed | Yes<br><br>(Involvement of sponsor in trial committee, sponsor owns choice of PRO) | Some concerns |
| Mereles et al 2006 | Permuted block randomization | No information provided | Investigators blinded | Description of withdrawals & method of imputation for missing data | All expected outcomes are reported | Yes<br><br>(73.3% PAH in control group, 86.6% PAH in treatment group, small sample size) | Some concerns |
| Chan et al 2013 | Randomised but not described how | No information provided | Investigator blind | Description of withdrawals & method of imputation for missing data | All expected outcomes are reported | Yes<br><br>(Small sample size, only female participants) | High |
| Weinstein et al<br><br>2013 | Randomised using 2 x 2 sequential block randomization schema of subject numbers | No information provided | Investigators blinded | Description of withdrawals & method of imputation for missing data | All expected outcomes are reported | Yes<br><br>(Small sample size, all participants female) | High |

| | | | | | | | |
|---|---|---|---|---|---|---|---|
| Saglam et al 2015 | Randomised through a program to generate random numbers | No information provided | No information provided | Description of withdrawals | All expected outcomes are reported | Yes (Small sample size) | Some concerns |
| GALILEO-PAH 2017 | Randomised but not described how | No information provided | Not blinded | Description of withdrawals & method of imputation for missing data | All expected outcomes are reported | Yes (Small sample size, open-label) | High |
| Babu et al 2019 | Randomised but not described how | No information provided | Not blinded | Description of withdrawals & method of imputation for missing data | All expected outcomes are reported | Yes (Only 40.5% of patients PAH) | High |
| Karapolat et al 2019 | Randomised using computer programme | No information provided | Single-blind | Description of withdrawals & method of imputation for missing data | All expected outcomes are reported | Yes (Small sample size) | Medium |
| Aslan et al 2020 | Randomised using computer-based random number generator | Allocation concealment through prefilled envelopes | Single-blind | Description of withdrawals & method of imputation for missing data | All expected outcomes are reported | Yes (66.7% PAH in TIMT group, 83.3% PAH in Sham group, small sample size) | Some concern |
| Yilmaz et al 2020 | Randomised using computer-based block randomisation | No information provided | Double blind | Description of withdrawals & method of imputation for missing data | All expected outcomes are reported | Yes | Low |
| EU-TRAIN-01 2021 | Randomised using permuted block randomization procedure, sealed envelopes, stratified by centre | Concealment using sealed envelopes | Investigators involved in data analysis blinded, participants not blinded | Description of withdrawals & method of imputation for missing data | All expected outcomes are reported | Yes | Some concerns |
| Hemnes et al 2021 | Randomised, stratified by sex and functional class | No information provided | Single blind | Description of withdrawals & method of imputation for missing data | All expected outcomes are reported | No | Some concerns |

| | | | | | | | |
|---|---|---|---|---|---|---|---|
| Kahraman et al 2023 | Block randomisation using random block sizes of 2 | Conducted by an independent party | Evaluator blind | Description of withdrawals & method of imputation for missing data | All expected outcomes are reported | Yes<br><br>(83.3% / 91.7% PAH patient, small sample size) | Some concerns |

*Supplementary table E4: GRADE of Evidence*

| **Number of Studies**<br><br>**(Number of Participants)** | **HRQoL Assessment**<br><br>**(Number of Participants, studies)** | **Quality Assessment** | | | | | **Quality** |
|---|---|---|---|---|---|---|---|
| | | **Study Limitations** | **Consistency** | **Directness** | **Precision** | **Publication Bias** | |
| **Combination** | | | | | | | |
| 4 (360) | SF-36 (267,1)<br><br>EuroQoL (40,1)<br><br>MLWH (27,1)<br><br>QoL score (26,1) | Limitations in some studies<br><br>(-1) | Consistent | Direct | No important imprecision | Unlikely | +++, moderate |
| **ERA** | | | | | | | |
| 6 (1104) | SF-36 (1007,4)<br><br>Subject global assessment (64,1)<br><br>DFI (33,1) | No significant limitation | Consistent | Direct | No important imprecision | Unlikely | ++++, high |
| **Exercise** | | | | | | | |
| 13 (580) | SF-36 (382,8)<br><br>emPHasis10 (42,1)<br><br>BDI (30,1)<br><br>IPAH (30,1)<br><br>MLHF (30,1) | Limitations in some studies<br><br>(-1) | Inconsistent<br><br>(-1) | Direct | No important imprecision | Unlikely | ++, low |

| | | | | | | | |
|---|---|---|---|---|---|---|---|
| | NHP (53,2)<br>FSS (28,1)<br>HAP (28,1)<br>CAMPHOR (23,1)<br>LPHQ (22,1) | | | | | | |
| **IP** | | | | | | | |
| 108 (1) | PAH-SYMPACT (108,1) | No significant limitation | Only 1 study<br>(-1) | Direct | No important imprecision | Unlikely | +++, moderate |
| **Novel** | | | | | | | |
| 323 (1) | PAH-SYMPACT (323,1) | Limitations in study<br>(-1) | Only 1 study<br>(-1) | Direct | No important imprecision | Unlikely | ++, low |
| **PDE5i** | | | | | | | |
| 405 (1) | SF-36 (405,1)<br>EQ-5D (405,1) | No significant limitation | Only 1 study<br>(-1) | Direct | No important imprecision | Unlikely | +++, moderate |
| **Prostanoids** | | | | | | | |
| 1917 (9) | MLWHF (821,3)<br>DFI (851,4)<br>EuroQoL (203,1)<br>TSQM-9 (42,1)<br>Chronic HF (20,1)<br>NHP (20,1) | No significant limitation | Inconsistent<br>(-1) | Direct | No important imprecision | Unlikely | +++, moderate |
| **sGCS** | | | | | | | |

| | | | | | | | |
|---|---|---|---|---|---|---|---|
| 839 (2) | LPHQ (839,2)<br><br>EQ-5D (396,1) | No significant limitation | Inconsistent<br><br>(-1) | Direct | No important imprecision | Unlikely | +++, moderate |
| **Other** | | | | | | | |
| 354 (9) | CAMPHOR (91,3)<br><br>MLHFQ (41,2)<br><br>SF-36 (191,3)<br><br>emPHasis-10 (150,1)<br><br>PGA (56,1)<br><br>IPAQ-SF (31,1)<br><br>McMaster Chronic Respiratory Questionnaire-Dyspnoea subscale (23,1)<br><br>Self-rated breathlessness score (23,1)<br><br>Self-rated quality of sleep (23,1) | Limitations in some studies<br><br>(-1) | Consistent | Direct | No important imprecision | Unlikely | +++, moderate |

Supplementary figure E2: inclusion (A) and search strategy (B)

A

**Included:**

PROMs identified from systematic review with MCID in PAH powered for HRQoL outcome [these were LPHQ (or MLWHF-PH), SF-36, EQ-5D-5L]
PROMs powered for HRQoL outcome in forthcoming RCTs with a MCID in PAH registered on ClinicalTrials.gov [EmPHasis-10]
Pulmonary Hypertension (group 1 (PAH) and group 4 (CTEPH))

**Excluded:**

Studies without measurement property evaluation or insufficient detail for analysis (e.g. abstracts)
Condition-specific PROMs (CAMPHOR, PAH-SYMPACT) that fail to meet inclusion criteria (reason: objective is to draw meaningful conclusions on HRQoL outcomes in historical, or forthcoming RCTs.)
MLWHF that is not PH (reason: as this is not condition specific)
Psychometric studies with included PROMs of conditions other than group 1 or group 4 pulmonary hypertension (reason: not population of interest)

B

**PubMed 1st Jan 2001 to 1st May 2024**
Search: ((pulmonary hypertension) AND (EQ-5D-5L OR EuroQoL OR EmPHasis-10 OR Living with Pulmonary Hypertension OR LPH OR MLWHF OR Minnesota Living with Heart Failure)) AND (validation OR psychometric) Sort by: Most Recent
("hypertension, pulmonary"[MeSH Terms] OR ("hypertension"[All Fields] AND "pulmonary"[All Fields]) OR "pulmonary hypertension"[All Fields] OR ("pulmonary"[All Fields] AND "hypertension"[All Fields])) AND ("EQ-5D-5L"[All Fields] OR "EuroQoL"[All Fields] OR "EmPHasis-10"[All Fields] OR (("lived"[All Fields] OR "lives"[All Fields] OR "living"[All Fields] OR "livings"[All Fields]) AND ("hypertension, pulmonary"[MeSH Terms] OR ("hypertension"[All Fields] AND "pulmonary"[All Fields]) OR "pulmonary hypertension"[All Fields] OR ("pulmonary"[All Fields] AND "hypertension"[All Fields]))) OR "LPH"[All Fields] OR "MLWHF"[All Fields] OR (("minnesota"[MeSH Terms] OR "minnesota"[All Fields] OR "minnesota s"[All Fields]) AND ("lived"[All Fields] OR "lives"[All Fields] OR "living"[All Fields] OR "livings"[All Fields]) AND ("heart failure"[MeSH Terms] OR ("heart"[All Fields] AND "failure"[All Fields]) OR "heart failure"[All Fields]))) AND ("valid"[All Fields] OR "validate"[All Fields] OR "validated"[All Fields] OR "validates"[All Fields] OR "validating"[All Fields] OR "validation"[All Fields] OR "validational"[All Fields] OR "validations"[All Fields] OR "validator"[All Fields] OR "validators"[All Fields] OR "validities"[All Fields] OR "validity"[All Fields] OR ("psychometrical"[All Fields] OR "psychometrically"[All Fields] OR "psychometrics"[MeSH Terms] OR "psychometrics"[All Fields] OR "psychometric"[All Fields]))

**Cochrane Search 'all dates'**
(pulmonary AND hypertension):ti,ab,kw AND (Emphasis-10 OR SF-36 OR EQ-5D-5L OR EuroQoL OR Living with Pulmonary Hypertension OR MLWHF) AND (psychometric OR validation OR quality of life) NOT (neonates OR child) (Word variations have been searched)
Scopus – systematic review and citation searching (pulmonary AND hypertension) ti,ab,kw: AND (psychometric) ti,ab,kw AND (review) ti,ab,kw

*Supplementary figure E3 PRISMA-COSMIN flow chart (adapted)*

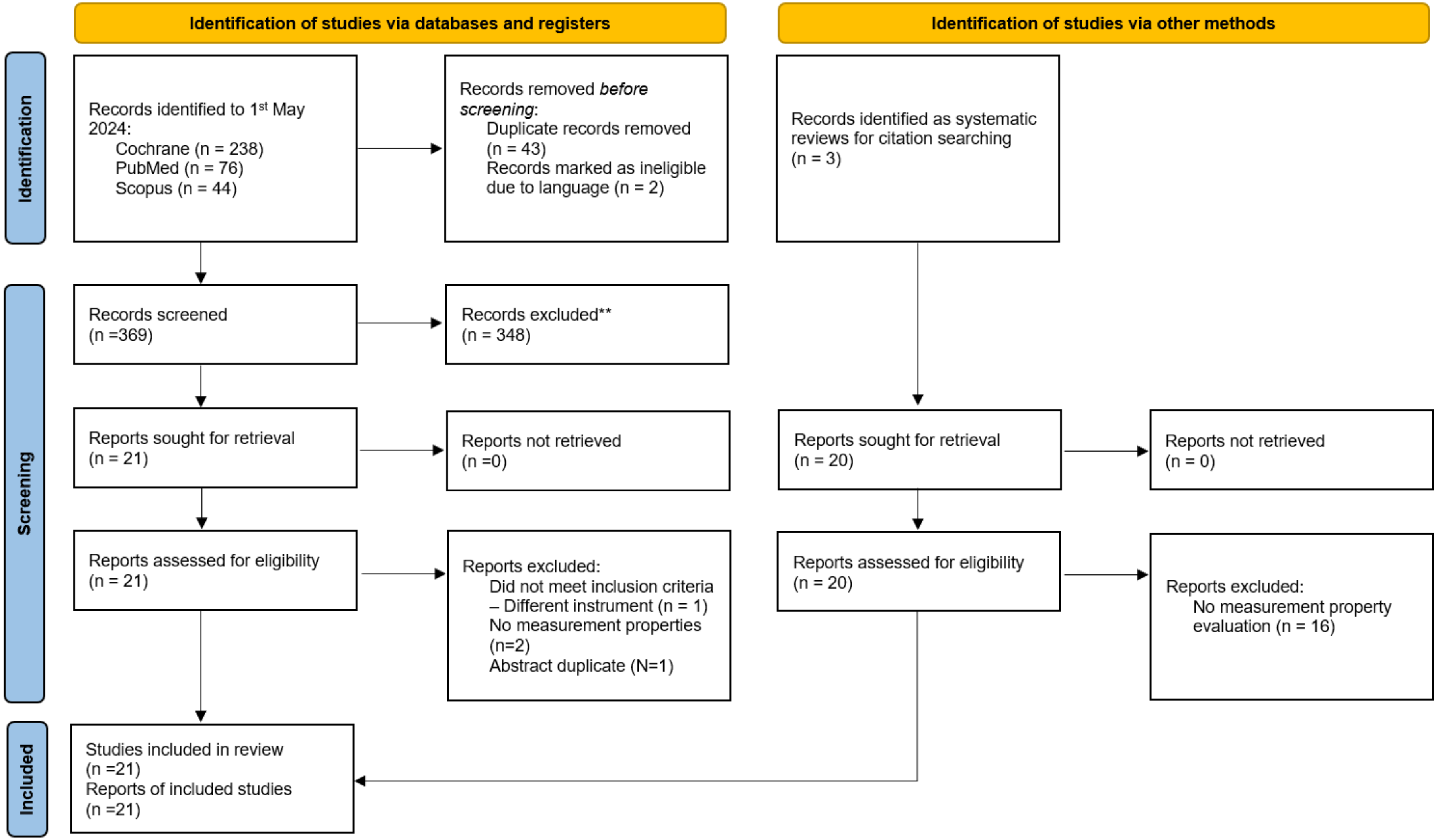

*Supplementary table E5 Definition of terms outlined within COSMIN criteria of assessment of measurement properties*

| | Definition of terms outlined within COSMIN assessment of measurement properties | |
|---|---|---|
| PROM Design | Patient reported outcome measure (PROM) | An instrument validated to capture subjective reporting of the individual experience of a health condition that measures changes in health-related quality of life that are important to the disease population. |
| | Concept elicitation | Qualitative coding of 'concepts of interest' by at least two researchers independently. Each item should be tested for comprehensibility. |
| | Content validity and interpretability | How well the PROM covers all relevant aspects of HRQoL; includes saturation analyses. Includes comprehensibility, which should be checked by patients and professionals to determine overall interpretability. Interview sample size should be ≥7 or via survey ≥50. |
| Internal Structure | Structural validity | Statistical analysis performed in an appropriate sample size based on the type of psychometric analysis e.g. confirmatory factor analysis |
| | Internal consistency | Reliability within the PROM using different sets of items. An appropriate statistical method (e.g. Cronbach's alpha) is chosen to determine the response agreement (adequate ≥0.7) between items |
| | measurement invariance\cross-cultural validity | Multi-group factor analysis or differential item functioning performed to assess for differences (age, gender, language) to determine whether PROM performs well across all groups. |
| Variability and Error | Test-retest, inter-\intra-rater reliability | Test-retest evaluates performance under stable conditions. Different responses on same occasion (inter-) or by the same person on different occasions (intra-) determine responder reliability. The intraclass correlation coefficient (ICC) ) is accepted as >0.7. |
| | Instrument measurement error and *response shift* | Systematic and random error in scores that is not attributed to true changes in a construct should be reported.<br>*Response shift* is whereby an individual may reprioritise certain HRQoL values rather than experience a true change in HRQoL.(58) In PAH, this may depend on time in the disease course.(59,60) |
| PROM Hypothesis Testing | Criterion validity | The extent to which the PROM agrees with a 'gold standard' using statistical testing (e.g. AUC or sensitivity) - agreed by expert consensus. E.g. a physical item and correlation with 6MWD |
| | Construct and Convergent validity | Assuming the PROM is valid, construct validity considers how well scores correlate with a hypothesis (e.g. WHO FC groups). Convergent validity is correlation with PROMs measuring similar constructs is. |
| | Discriminative validity | Statistical comparison of performance of PROM between known subgroups. For example, risk classification (e.g. COMPERA 2.0) |
| | Responsiveness | Ability of the PROM to detect change over time in HRQoL. Can use either criterion approach: comparison to a gold standard, or construct approach: comparison before and after an intervention |

Table 1: Definition of terms outlined within COSMIN criteria of assessment of measurement properties.

Description of 4 stage approach to PROM assessment; design, internal structure, variability and error, and hypothesis testing. Individual criteria are summarised. Interventions are important for hypothesis testing. *6MWD six-minute walk distance; AUC area under the curve; COSMIN consensus-based standards for the selection of health measurement instruments; WHO FC World Health Organization Functional Classification*

*Supplementary Table E6 Interviews and surveys included for analysis of impact of PAH on HRQoL.*

| Reference | N= | Country | Demographics | Type | Acquisition |
|---|---|---|---|---|---|
| Peloquin et al (1998)(61) | 3 | Canada | 100% female<br>Age 24-37 | Semi-structured interview | Impact case-narrative |
| ¥Flattery et al (2005) (62) | 11 | USA | 63% female<br>Age 58.5 (40-72) | Semi-structured interview | Colaizzi's 7 step process |
| Taichman et al (2005)(63) | 155 | USA | 81% female<br>Age 53 years (18-84)<br>58% <3y treatment<br>63% iPAH<br>68% Caucasian<br>94% WHO FC II/III | Cross-sectional survey | SF-36, St George's Respiratory questionnaire incl. symptoms, haemodynamics, walk-distance |
| ¥McDonough et al (2010)(64) | 10 | USA | 70% female<br>Age 65 (38-81)<br>80% Caucasian | Semi-structured interview | NVIVO 8.0 for thematic analysis |
| Studer and Migliore (2011)(65) | 19 | USA | Majority group 1 PAH<br>Age 35-75 years<br>WHO FC II-III | Semi-structured interview Patients (19) and caregivers (12) | |
| Guillevin et al (2013)(66) | 455 | Europe<br>(5 countries) | 74% female<br>Age 52 | Survey – patients (326) and caregivers (129) | Scoping interviews, paper questionnaires |
| ¥Kingman et al (2014)(59) | 39 | 7 Countries Globally | 74% female<br>% PAH<br>Age range 19-91<br>79% WHO FC II/III | Ethnography | 140 hours observational video transcribed |
| ¥Yorke et al (2014)(67) | 30 | UK | 60% female<br>Age 56.3 (26-80)<br>40% iPAH<br>63% diagnosis ≥5years | Semi-structured interview | Thematic analysis |
| Zhai et al (2017)(68) | 174 | China | 81% female<br>Age 32.9<br>61% CHD-PAH<br>93% WHO FC II/III | Survey – patients (114) & caregivers (60) | Scoping interviews, paper questionnaires |
| Armstrong et al (2019)(69) | 567 | UK | 70% female<br>Age 59 | Survey - patients | PPI design, 11% online, 89% paper |
| Rawlings et al (2020)(70) | >1900 | Europe, Asia, Americas | NA | Qualitative systematic review | Thematic synthesis |
| PHA UK (2021)(71) | 119 | UK | 92% female<br>77% Age 31-70<br>72% PAH | Survey - patients | 100% online |
| PHA UK (2022)(72) | 112 | UK | 77% female<br>Age 45-65<br>91% Caucasian<br>65% diagnosis ≥3years | Survey - patients | 100% online<br>(65% mobile phone) |
| Tremblay et al (2022)(73) | 257 | Canada | 83% female<br>Age 42% ≥65y<br>89% Caucasian (European)<br>42% iPAH<br>53% WHO FC II | Questionnaire (Likert-scale) | Categorical/numerical analysis |
| PHA UK (2023)(74) | 859 | UK | 73% female<br>Age 64<br>93% white | Survey - patients | 66% paper<br>(34% online) |
| PH Global Patient Survey*(2024)(75) | 3335 (adults) | Global<br>(88 countries) | 81.6% female<br>Age 52.1y<br>59.6% Group 1 PAH | Survey - patients (2890) & caregivers (445) | 100% online with descriptive analysis of categorical/numerical data & thematic analysis of free-text |

Age in years, median and range where available. CHD-PAH congenital heart disease pulmonary arterial hypertension, iPAH idiopathic pulmonary arterial hypertension, WHO FC World Health Organisation Functional Class. (n=8045) ¥Papers included in qualitative systematic review reviewed for narrative agreement by FV/RB prior to full synthesis of published systematic review (70) *formal results unpublished at time of this manuscript submission(75)

#### Supplementary table E7 Quality of PROM design and development

| PROM | | PROM design | | | | | | | Cognitive interview (CI) study[2] | | | TOTAL PROM DEVELOPMENT |
|---|---|---|---|---|---|---|---|---|---|---|---|---|
| | | General design requirements | | | | | Concept elicitation[1] | Total PROM design | General design requirements | Comprehensibility | Total CI study | |
| | Language in which the PROM was developed | Clear construct | Clear origin of construct | Clear target population for which the PROM was developed | Clear context of use | PROM developed in sample representing the target population | | | CI study performed in sample representing the target population | | | |
| Emphasis-10(76) | English | V | V | V | V | V | I | I | V | I | I | I |
| LPHQ(8) | English, French, German | V | V | V | V | V | D | D | V | A | D | D |
| SF-36(63) | English | I | I | I | I | I | I | I | I | I | I | I |
| EQ-5D-5L | English | NA | NA | NA | NA | NA | NA | NA | NA | NA | NA | NA |

*Cosmin risk of bias assessment*

V = very good
A = adequate
D = doubtful
I = inadequate
NA = not available

*Supplementary table E8: [A] Detailed measurement properties [COSMIN risk of bias (A to I) and modified GRADE of evidence] and [B] Summary table*

A

| | Instrument | Study (ref) (n=) | Risk of bias quality rating (A to I) | Measurement property and results reported | Measurement property rating individual \| overall | | Overall evidence quality* |
|---|---|---|---|---|---|---|---|
| | Structural validity | | | | | | |
| Internal structure | LPHQ (2013)/ MLWHF-PH (from 2006) | Bonner (2013)(8)<br>N= 196 | D | CFA. Overall fit for emotional and physical scores. Total score poor overall fit with only five items meeting 0.7 threshold. Specific CFI or root means not reported | ? | ? | Low |
| | EmPHasis-10 | Yorke (2014)(76) N=226<br><br>Rawlings (2024)(77) N=263 | A<br><br>V | Rasch analysis - no CFI, TLI, local independence/monotonicity reported, item chi-square fit statistic (residual ±2.5, p>0.05), overall fit not reported<br><br>CFA (n=132) three factor model, RMSEA <0.06, TLI 98.9% and SRMR <0.08 | ?<br><br>+ | ?<br><br>+ | Inconsistent (different structures) |
| | SF-36 | NA | | | | | NA |
| | Internal consistency | | | | | | |
| | LPHQ (2013)/ MLWHF-PH (from 2006) | Cenendese (2006)(78) N= 48<br><br>Bonner (2013)(8) N= 196 | V | Cronbach's alpha 0.88 to 0.92<br>Test-retest 1 week apart<br>Cronbach's alpha 0.87 to 0.92 | +<br><br>+ | + | High |
| | EmPHasis-10 | Yorke (2014)(76) N=226<br>Takeyasu (2020)(79) N=76<br>Shi (2022)(80) N=76<br>Odevoglu (2019)(81) N=101<br>Surace (2019)(82) N=100<br>Rawlings (2024)(77) N=263 | V | Cronbach's alpha 0.9<br>Cronbach's alpha 0.86 (Japanese)<br>Cronbach's alpha 0.95 (Chinese)<br>Cronbach's alpha 0.98 (Turkish)<br>Cronbach's alpha 0.93 (Italian)<br>Cronbach's alpha 0.89 | +<br>+<br>+<br>+<br>+<br>+ | + | High |
| | SF-36 | Twiss (2013)(83) N=65<br>Fernandes (2014) (84) N=54 | V<br>V | Cronbach's alpha 0.74 to 0.94<br>Cronbach's alpha 0.85 | +<br>+ | + | High |
| | Cross-cultural validity/measurement invariance | | | | | | |
| | LPHQ/MLWHF | No multi group factor analysis or differential item functioning (DIF) studies for measurement invariance across age/gender/time since diagnosis | | | - | - | NA |
| | EmPHasis-10 | Takeyasu (2020)(79) N=76<br>Lewis (2021)(85) N=1745<br>Hendriks (2021)(1) N=61<br>Shi (2022)(80) N=76 | I<br>I<br>I<br>I | Cronbach's alpha 0.86 (Japanese)<br>Cronbach's alpha 0.95 (Chinese)<br>Cronbach's alpha 0.98 (Turkish)<br>Cronbach's alpha 0.93 (Italian) | -<br>-<br>-<br>- | - | Very Low |

| | | | | | | | |
|---|---|---|---|---|---|---|---|
| | | Borgese (2021)(86) N=565 | I | (US) Older age associated with higher scores, females higher scores than males, higher BMI associated with higher scores<br>No MGFA for measurement invariance or DIF<br>SEM&RCI same for new treatment Vs diagnosed within 6 months and regardless of PAH aetiology. | - | | |
| | SF-36 | No studies for measurement invariance across age/gender/time since diagnosis | | | - | - | |
| | Test-retest reliability | | | | | | |
| | LPH (2013)/ MLWHF-PH (from 2006) | Cenendese (2006)(78) N=26 | I | Spearman's r=0.94 (total), physical r=0.93, emotional r=0.9 (p<0.001) | ? | ? | Very Low |
| | EmPHasis-10 | Yorke (2014)(76) N=33<br>Takeyasu (2020)(79) N=76<br>Odevoglu (2019)(81) N=101<br>Surace (2019)(82) N=100<br>Borgese (2021)(86) N=565 | A<br>A<br>A<br>I<br>A | ICC 0.95<br>ICC 0.86<br>ICC 0.97<br>t-test correlation<br>ICC 0.95 | + | + | High |
| | SF-36 | Twiss (2013)(83) N=65<br><br>Gilbert (2009)(10) N= 207 | V<br><br>I | Test-retest correlation (spearman's) (%$r^2$)<br>Physical 0.93(86%), RP 0.81 (66%), pain 0.72(52%), GH 0.94(88%), vitality 0.78(61%), SF 0.76(58%), RE 0.7(49%), MH 0.7(56%)<br><br>Stated ICC calculated between screening and baseline but values not reported | ?<br><br>- | ? | Moderate |
| | Measurement Error | | | | | | |
| | LPHQ (2013)/ MLWHF-PH (from 2006) | Bonner (2013)(8) N= 196 | I - No SDC | MID: LPHQ Emotional Score 1.48-3.69, Physical Score 1.88 to 4.71, LPHQ Total score 4.41 to 11.02 | - | - | Very Low |
| | EmPHasis-10 | Lewis (2021)(85) N=1068<br><br>Hendriks (2021)(1) N=61 | I - No SDC<br><br>V | MDC = 9 (n=1068, 61%) based on field walk distance. Sensitive in incident, not prevalent patients. Improvement from high risk -12 (-6 to -19), deterioration to high risk +13 (8-17)<br>Anchor MCID -9, distributional -6 (mean -8)<br><br>SEM (-5), ES (-6). Consistent for subtypes of PAH, SDC = 1 for 6MWD and =3 for WHO FC | -<br><br>+ | + | Moderate - inconsistencies |

| | | | | | | | |
|---|---|---|---|---|---|---|---|
| | SF-36 | Twiss (2013)(83) N=65 | V | SEM physical = 6, RP = 12, pain = 13, GH = 5, vitality = 11, SF = 15, RE = 17, MH = 9. low levels of explained variance ($r^2$<0.7) except physical and GH | ? | + | High |
| | | Gilbert (2009)(10) N= 207 | V | For moderate ES: MID (SEM); physical 13 (9), RP = 25 (16), SF = 21 (15), vitality 15 (10) | + | + | |
| | Criterion validity | | | | | | |
| | LPHQ/MLWHF-PH | NA - No gold standard for sensitivity/specificity | | | | | |
| | EmPHasis-10 | NA - No gold standard for sensitivity/specificity | | | | | |
| | SF-36 | NA - No gold standard for sensitivity/specificity | | | | | |
| | Comparison with other instruments of similar construct | | | | | | |
| | LPHQ (2013)/ MLWHF-PH (from 2006) | Cenendese (2006)(78) N= 48 | A | SF-36 (r=0.62) | + | + | Moderate - indirect |
| | | Bonner (2013)(8) N= 190 | A | LPHQ emotional EQ-5D anxiety/depression (0.59) and self-care (0.24), LPH physical EQ-5D mobility (0.51), usual activities (0.46), anxiety/depression (0.42), Total and EQ-5D index 0.57. | + | | |
| | EmPHasis-10 | Yorke (2014)(76) N=226 | V | HAD (r=0.77), MLQHF-PH (r=0.61) | + | + | High |
| | | Takeyasu (2020)(79) N=76 | V | MLWHF-PH (r=0.77), HAD total (0.52), EQ-5D (-0.64), English version (r=0.61), Turkish (r=0.85) | + | | |
| | | Shi (2022)(80) N=76 | A | PCS (r=-0.85), MCS (r=-0.82), EQ-5D (r=-0.72, p=0.002) | + | | |
| | | Odevoglu (2019)(81) N=76 | V | MLWHF (r=0.85, p=0.001) | + | | |
| | | Surace (2019)(82) N=100 | D | SF-36 and NHP item-item correlation | ? | | |
| | SF-36 | Gomberg-Maitland (2008)(87) N=147 | V | Correlation with CAMPHOR scales, >0.7. Pain scale lowest overall correlation, r=0.51. | + | + | High |
| | Response to intervention | | | | | | |
| | LPHQ (2013)/ MLWHF-PH (from 2006) | Cenendese (2006)(78) N= 38 | A | Response to vasodilator (-10) p<0.001 ES -0.48 with 6MWD 50m. Better outcome with score <40. | + | + | Moderate |
| | | Bonner (2013)(8) N= 196 | D | Phase III clinical trial intervention not detailed. No ES by therapy, response to change in WHO FC/Borg/6MWD, no anchor | - | | |

| | | | | | | | |
|---|---|---|---|---|---|---|---|
| | | Chua (2006)(88) N=83 | D | Treprostinil, Bosentan – samples all pooled with paired time points in relation to 6MWT and NYHA class (r>-0.5, p<0.01) | - | | |
| | | PATENT-1(89) N=224 (2.5mg)<br>*Placebo Vs riociguat* N=55 (1.5mg)<br>MID Total score -4 to -11 | V<br>6MWD 36m | Overall difference at 8 weeks -6 (-10 to -3), p=0.002, week 12 for 2.5mg dose (-7.6±16.8), week 12 for 1.5mg dose (-11.8±16.1). Average change for placebo -2.7±15.1 | + | | |
| | | PATENT-2 (90) N=209 (2.5mg)<br>*Placebo Vs riociguat* N=54 (1.5mg) | V<br>6MWD 51m | 0 and 12 weeks. Change at week 12, 2.5mg dose (-11.0±18.9), 1.5mg dose (-14.3±18.9). Average change for placebo -2.1±15.7 | + | | |
| | | TRIUMPH-I(46) N=235 | V | | | | |
| | EmPHasis-10 | NA | | | | | |
| | SF-36 | Chua (2006)(88) N=21 | D | Sildenafil & 6MWD correlates with items: r=0.68 physical function, r=0.65 RP, r=0.59 RE r=0.62 pain (p<0.01) baseline and 3 months | ? | + | High |
| | | Gilbert (2009)(10) N= 207 | A | Sildenafil Vs placebo. Placebo effects for physical, RP, SF, vitality. Statistical significance by $\chi^2$ observed only for the physical item (p = 0.034) | + | | |
| | | EARLY(2008)(47) N=74<br>*Placebo Vs bosentan* | V | 57% bosentan, 43% placebo: Physical, RP, pain, GH, vitality, SF, MH, RE all p>0.05 | + | | |
| | | PACES(2008)(49) N=267<br>*Placebo Vs sildenafil on IV epoprostenol*<br>*MCID for moderate ES (SEM); physical 13 (9), RP = 25 (16), SF = 21 (15), vitality 15 (10)* | V<br>6MWD<br>+28.8m | Placebo Vs sildenafil (all <MID); for physical (0.3[CI,4.7 to 4.1] vs. 7.8 [CI, 3.6 to 12.1];P0.003), GH (1.4 [CI,4.8 to 2.1] vs. 6.6 [CI, 3.3 to9.9];P0.001), vitality (0.8 [CI,3.3 to 4.9] vs. 10.2[CI, 6.2 to 14.2];P0.001), social functioning (2.5[CI,7.8 to 2.9] vs. 4.0 [CI,1.1 to 9.2];P0.049),and mental health (3.7 [CI,7.0 to0.5] vs. 3.0 [CI,0.1 to 6.2];P0.001), but not RE, RP, bodily pain p>0.05 | + | | |
| | | ARIES2(2008)(48) N=394<br>*Placebo Vs ambrisentan (5mg dose)* | A<br>6MWD<br>+45-59m | Placebo Vs ambrisentan (<MID); Physical (-0.2±7.14) vs (3.41±6.96) p=0.005. Others not reported | + | | |

| | | | | | | | | |
|---|---|---|---|---|---|---|---|---|
| | | EU-TRAIN-01)2021)(18)<br>*Exercise intervention* | N=116 | V<br>6MWD<br>34±8.3m | Change RP score 20.6±38.6, p=0.07, physical 8.1±18.1, p=0.23, all other items p>0.05 other than MH 5.2±12.5, p=0.004 (no MID) | + | | |
| | EQ-5D-5L<br><br>*MCID ILD*(91)<br>*6MWD 35m*<br>*Anchor 0.017 (index range -0.148 to 0.949). Index range for PH WHO FC I 0.274 to 0.93 for WHO FC IV*(92) | AIR (2002)(56)<br>*Placebo Vs Inhaled Iloprost*<br><br>PATENT-1(2013)(34)<br><br><br><br><br>PATENT-2(2015)(35) | N=203<br><br><br>N=227 (2.5mg)<br>N=55 (1.5mg)<br>N=108 (placebo)<br><br><br>N=325 | V<br>6MWD 36m<br><br>V<br>6MWD 36m<br><br><br><br>V<br>6MWD 51m | Health state score improved by 0.09 in iloprost group (p<0.05)<br><br>2.5mg Vs placebo change at 8 weeks from baseline -0.06(0.01-0.11), p=0.07; week 12 2.5mg dose (0.05±0.21), 1.5mg (0.10±0.26), placebo 0.02±0.22<br><br>2.5mg dose at week 12 (0.06±0.22), 1 year (0.06±0.24); 1.5mg dose week 12 (0.15±0.24), 1 year (0.13±0.24); placebo week 12 (0.03±0.24), 1 year (0.07±0.20) | + | + | High |
| | Subgroup and known-group analyses (e.g. sex, age, illness duration, WHO FC) | | | | | | | |
| | LPHQ (2013)/ MLWHF-PH (from 2006) | Chua (2006)(88)<br><br><br><br>Cenendese (2006)(78)<br><br><br>Zlupko (2008)(93)<br><br><br><br><br><br>Bonner (2013)(8) | N= 21<br><br><br><br>N= 48<br><br><br>N= 93<br><br><br><br><br><br>N= 196 | V linear mixed effects model<br><br><br>V<br><br><br>A<br><br><br><br><br><br>A | NYHA FC total 13.87 ±2.54 (p<0.0001). Rate of change per 1 metre 6MWT -0.11±0.02(p<0.0001) No haemodynamic association.<br><br>Physical Score with cardiac index (r=-0.48) SvO2 (r=-0.59), total score &WHO FC 0.61 (<0.001)<br><br>Poor overall haemodynamic correlation. RA pressure and physical score (0.29, p=0.05), total 0.36, p=0.01). WHO FC II Vs III emotional (p>0.05), physical (p<0.005)<br>WHO FC II, III & IV, the total score was 27 (95% CI 9–45), 41 (34–48) and 62 (55–69), respectively (p <0.001 for class III *versus* IV). | +<br><br><br><br>+<br><br><br>?<br><br><br><br><br>? | | High |
| | EmPHasis-10 | Yorke (2014)(76)<br><br><br>Takeyasu (2020)(79) | N=226<br><br><br>N=76 | A<br><br><br>V | WHO FC II and III (mean difference 10.9, 95% CI 7.3–14.5; p<0.001)<br><br>6MWD (r=-0.38), WHO FC (Jonckheere-Terpstra 1 sided, p<0.001) | ?<br><br><br>+ | + | High |

| | | | | | | | |
|---|---|---|---|---|---|---|---|
| | | Lewis (2021)(85) N=653 (6MWD) N=797 (ISWD) | A | 6MWD (r=-0.55, p<0.001), ISWD (r=-0.5,p<0.001), | + | | |
| | | Hendriks (2021)(1) N=61 | V | WHO FC (r=0.50) (6MWD, r=0.55; ISWD, r=0.50), haemodynamics (r=0.17–0.21). NYHA FC (variable 0,6,12mo), 6MWD (all timepoints, r=0.4-0.55, p<0.05), non-invasive risk score –(all time points, r<-0.4) | + | | |
| | | Shi (2022)(80) N=76 | D | PVR (r=0.46, p<0.001) | ? | | |
| | | Odevoglu (2019)(81) N=101 | A | WHO FC I (9.3±7.7), WHO FC II (12.7±9.9), WHO FC III (18.4±11.6) p=0.01, 6MWD r=0.29, p=0.003 | ? | | |
| | | Borgese (2021)(86) N=565 | V | Each 30-m increase in 6MWD was associated with a 1.3-point decrease in score (95% CI −1.6 to −1.0 point; p<0.001) β-coefficient for WHO functional class was 3.0 (95% CI 1.9–4.0; p<0.001). 6MWD explains 19% variance and WHO FC 21% | + | | |
| | | Favoccia (2024)(94) N=687 | A | Mortality - HR per 10-unit increase 1.27, 95% CI 1.03–1.56, *P* = 0.0 | + | | |
| | | Rawlings (2024)(77) N=263 | V | No correlation with years since PH diagnosis, WHO FC & fatigue (r=0.24) & breathlessness (r=0.22) | + | | |
| | SF-36 | Taichman (2005)(63) N=155 | A | 6MWD & PCS r=0.62 p<0.001, MCS r=0.33, p=0.06. PCS age r= -0.148 (p = 0.08) and MCS age r= 0.05 (p = 0.58). No correlation with haemodynamics. | + | ? Items vary | Moderate - inconsiste ncies |
| | | Chua (2006)(88) N=21 | V linear mixed effects model | Change in NYHA FC: physical -16.37 ±4.34 (p<0.002), RE -22.58±10.52 (p<0.05), pain 21.93±7.35(<0.01). Rate of change per 1 metre 6MWT: physical 0.12 ±0.03 (p<0.003), RE 0.24±-0.07(p<0.003)), no haemodynamic association. | + | | |
| | | Twiss (2013)(83) N=65 | A | WHO FC II Vs III PCS (p<0.05) and physical functioning (p<0.005) item only | ? | | |

| | | | | | | |
|---|---|---|---|---|---|---|
| | | Fernandes (2014)(84) | N= 54 | D | Baseline correlation PCS 6MWD (r = 0.493, p < 0.01) and WhO FC (r = -0.576, p < 0.01) | ? |
| | | Shi (2022)(80) | N=76 | A | P<0.001, WHO FC for I/II Vs III/IV for physical, and vitality & RE single not reproducible, other domains not significant. PCS and PVR (WU) r=-0.39 (p<0.001), MCS r=-0.31(p<0.001) | + |
| | | Li(2022)(95) | N=559 | D | PVR & PCS r=-0.15, p=0.005, not with MCS. | + |

%$r^2$ =% of explained variance; 6MWT six-minute walk test; AQOL Australian Quality of Life; CFA confirmatory factor analysis; CFI comparative fit index; DIF differential item functioning; EF energy fatigue, ES effect size; GH general health; HR hazard ratio; ICC intraclass correlation coefficient (adequate ≥0.7); ILD interstitial lung disease; MCID minimal clinically important difference; MCS mental component score, MGFA multi-group factor analysis; MH mental health, MID minimal important difference; NYHA New York Heart Association, r= correlation coefficient by Pearson or Spearman-Rank, interpreted as 0.1 weak, 0.3 moderate, 0.5 strong. PCS physical component score, PVR pulmonary vascular resistance, RCI reliable change index; RMSEA root mean square error of approximation, RE role emotional, RP role physical, SDC smallest detectable change (within patient group), SEM standard error of measurement (calculated as standard deviation ($\delta$) and reliability (r): SEM=$\delta\sqrt{(1-r)}$) can come from a single study, SF social functioning, SRMR standardised root mean square residual, TLI Tucker-Lewis Index, WHO FC World Health Organisation functional classification

B **SUMMARY Risk of bias assessment**: To determine the overall quality of a single study on a measurement property, the lowest rating is taken from V very good, A adequate, D doubtful, I inadequate. If one of the four criteria in the structural validity box is 'inadequate' then the overall methodological quality of structural validity is 'inadequate'.

***Modified GRADE approach** and risk of bias: quality of evidence as high, moderate, low or very low. Risk of bias assessed by COMIN risk of bias checklist. Risk of bias is downgrade by 1 level (serious) if multiple studies of doubtful quality and only one study of adequate quality, 2 levels (very serious) if multiple studies of inadequate quality, or only one study of doubtful quality or 3 levels (extremely serious) if only one study of inadequate quality available. Concerns considered in terms of inconsistency imprecision and indirectness. Indirectness not included as studies only in PAH meet inclusion criteria

| GRADE Factors | PROM | | |
|---|---|---|---|
| | Emphasis 10 | LPHQ | SF-36 |
| COSMIN Risk of Bias assessment | -2<br>*(No adequate studies, 2 D Grade)* | -2<br>(No adequate studies, 2 D Grade) | -3<br>(One inadequate study) |
| Inconsistency<br>*unexplained inconsistency of results across studies* | + | + | -1 |
| Imprecision (Sample Size) | + | + | + |
| Indirectness | n/a | n/a | n/a |
| OVERALL GRADE Quality Score | Low | Low | Very low |
| OVERALL COSMIN Recommendation | A | A | B |

*Supplementary table E9: Summary of PRISMA-COSMIN review*(96) *criteria*

| | Summary of PRISMA-COSMIN Criteria (2024) | Review meets criteria | Comments |
|---|---|---|---|
| | Title | +/- | NA – Journal title restrictions |
| | Abstract (Open science/background/methods/results/discussion) | +/- | Not all reported – multi-stage review with original research |
| | Plain Language Summary | + | |
| | Open Science (Registration/Conflict of interest / Support / Data availability, code and materials) | + | |
| Introduction and aims | Rational for review in context of existing knowledge | + | |
| | Elements included in research aim: | | |
| | Construct of interest | + | Background |
| | Population of interest | + | |
| | Type of measurement instrument of interest | + | |
| | Measurement properties of interest | + | |
| Methods | All available instruments included | +/- | |
| | Only instruments included that have at least some evidence of measurement properties | + | If valid in PH |
| | Search strategy described | + | |
| | Number of databases searched: | | Suppl E4 |
| | Searched in at least 2 databases | + | |
| | PUBMED/Cochrane | + | |
| | Additional databases – Scopus (citation searching) | + | |
| | Reference checking used | + | |
| | No time limits used or good arguments for time limit | + | |
| | No language restrictions used (google translate utilised) | + | |
| | Inclusion and exclusion criteria clearly described | + | Suppl E3(A) |
| | Reasons for excluding articles reported | + | Suppl E3(B) |
| | Abstracts and full-text article selection by at least 2 reviewers | +/- | Second author used in uncertainty |
| | Measurement property assessments extracted by at least 2 reviewers | + | Methods |
| | One of the authors of the review is also the developer of one of the instruments evaluated in the review | +/- | Authors did not perform analysis |
| Results | OMI (PROM) Characteristics: | | Table 2 |
| | Present characteristics of each included PROM | + | |
| | Present interpretability aspects for each included PROM | + | |
| | Present feasibility for each included PROM | + | |
| | Study Characteristics: cite each study report evaluating measurement properties and characteristics | + | Suppl E8 |
| | Risk of Bias: assessment presented for each included study | + | Suppl E7/8 |
| | Results of Individual studies: results for the measurement properties reported as raw data and rated against quality criteria | + | Supple E7/8 |
| | Results for multiple studies on the same instrument somehow combined (e.g. best evidence synthesis or pooling) | + | Best evidence synthesis |
| | Data synthesis was performed: | | Presented in Table 2 and Suppl E7 and E8 |
| | Per measurement property | + | |
| | For domains (reliability, validity, responsiveness) | + | |
| | For whole instrument | + | |
| | Certainty of evidence: assessment of certainty / confidence in body of evidence for each measurement property of PROM | + | |
| | All measurement properties reported | + | |
| | Make recommendations for suitable OMIs for a particular use | + | |
| Discussion | Provide general interpretation of results in context of other evidence | + | Strengths and limitations, conclusion, graphical abstract |
| | Discuss limitations of evidence included in review and review process | + | |
| | Discuss implications of results for practice, policy and future research | + | |

*Supplementary figure E4 Mapping PROMs to conceptual framework*

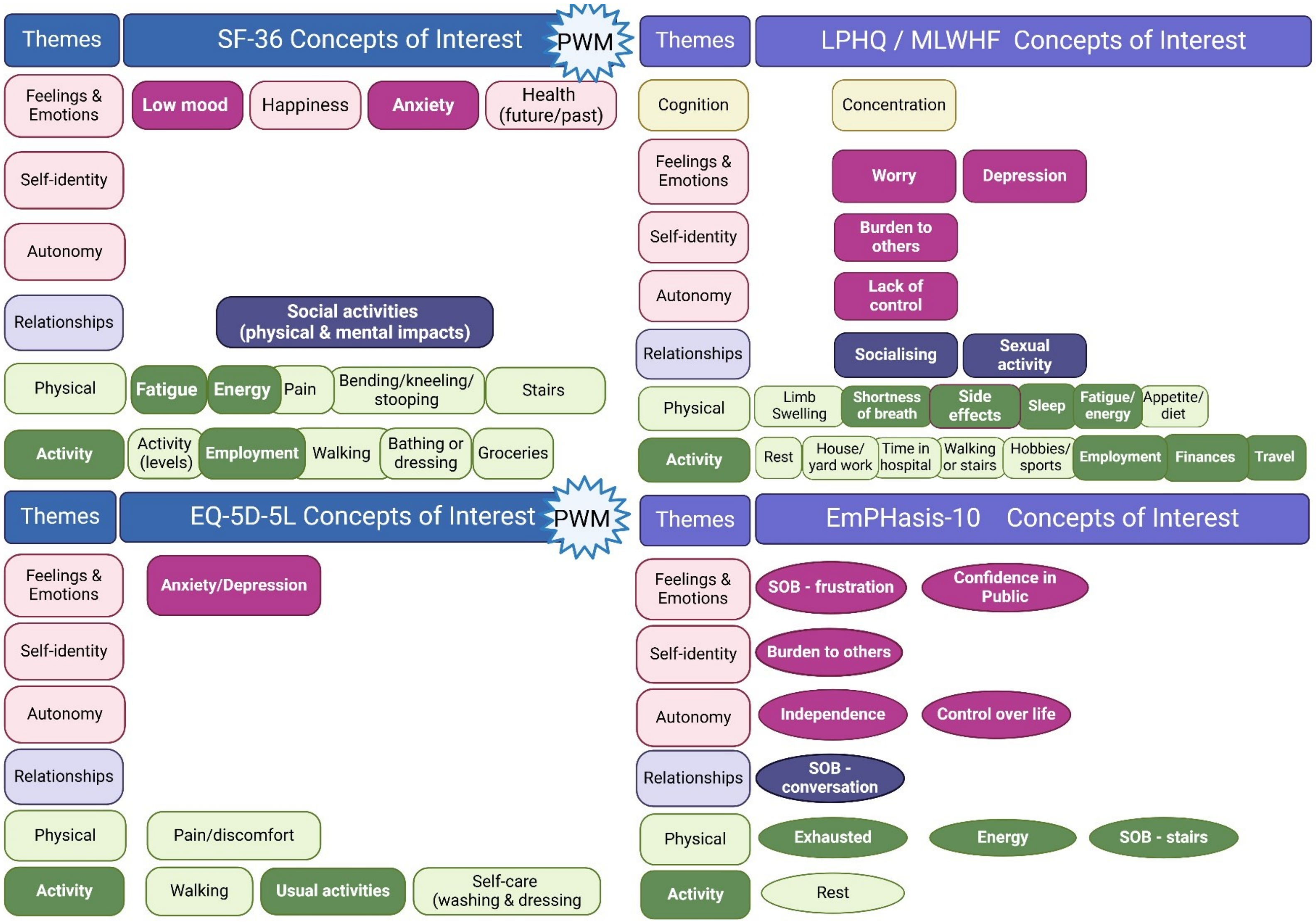

EQ-5D-5L EuroQol-5D-5L, LPHQ Living with Pulmonary Hypertension Questionnaire, MLWHF, Minnesota Living with Heart Failure Questionnaire, PWM preference weighted measure, SF-36 short form-36. Emphasis-10 (oval concepts) trials are underway for evaluation of responsiveness. https://BioRender.com/a50m191

32. van Campen JSJA, de Boer K, van de Veerdonk MC, van der Bruggen CEE, Allaart CP, Raijmakers PG, et al. Bisoprolol in idiopathic pulmonary arterial

hypertension: an explorative study. European Respiratory Journal. 2016 Sep;48(3):787–96.
